# COVID-19 Endemic Plan: Impact of Vaccination and Non-pharmaceutical Interventions with Viral Variants and Waning Immunity Using an Agent-Based Simulation

**DOI:** 10.1101/2022.06.03.22275834

**Authors:** Serin Lee, Zelda B. Zabinsky, Judith N. Wasserheit, Jennifer M. Ross, Shi Chen, Shan Liu

**Author notes:** Serin Lee, corresponding author.

## Abstract

**Backgrounds:** Despite the widespread distribution of SARS-CoV-2 vaccines, the COVID-19 pandemic continues with highly contagious variants and waning immunity. Low disease severity of the Omicron variant gives society hope that the COVID-19 pandemic could end.

**Methods:** We develop an agent-based simulation to explore the impact of COVID-19 vaccine willingness, booster vaccination schedule, vaccine effectiveness, and non-pharmaceutical interventions (NPIs) on reducing COVID-19 deaths while considering immunity duration and disease severity against the Omicron variant. The model is calibrated to the greater Seattle in year 2020 by observing local epidemic data. The simulation is run to the end of year 2024 to observe long-term effects.

**Results:** Results show that an NPI policy that maintains low levels of NPIs can reduce mortality by 35.1% compared to fully opening the society. A threshold NPI policy is especially helpful when the disease severity of the Omicron variant is high, or booster vaccines are not scheduled. A periodic booster schedule is needed to achieve the goal of lowering the number of deaths from COVID-19 to the level of influenza and pneumonia. Except for one scenario, 80% or more vaccine willingness is also needed to achieve this goal.

**Conclusions:** We find that a periodic booster vaccination schedule and mild disease severity of the Omicron variant play a crucial role in reducing deaths by the end of year 2024. If a booster schedule is not planned and the Omicron variant is not mild, NPI policies that limit society from fully opening are required to control the outbreak.

## 1. Introduction

SARS-CoV-2 vaccines authorized in the United States consistently show high effectiveness in reducing infections, hospitalizations, and deaths^1^. However, with the emergence of SARS-CoV-2 variants of concern (VOC) such as the Delta and the Omicron variant^2^ and uncertainty about the duration of both natural and vaccine-induced immunity, changes in the use of non-pharmaceutical interventions (NPIs), such as mask use, social distancing and school closures, and public reluctance to be vaccinated, the COVID-19 pandemic continues to ravage populations around the world. Questions remain whether the COVID-19 can become endemic and be treated like ‘seasonal influenza’. Understanding of the impact of vaccination and NPI policies on COVID-19 incidence, hospitalization, and death is needed to guide effective prevention and response effort.

The ongoing emergence of SARS-CoV-2 VOC has been a major threat so far. VOC may show increased transmissibility, disease severity, and/or resistance to immunity conferred by previous infection or vaccination^2^. The Alpha variant, which was detected first in the UK, and the Delta variant, which was first identified in India, has increased transmissibility although vaccine effectiveness against these variants seems to be similar to that against the original strain. The Omicron variant, first detected in South Africa, exhibits increased transmissibility and some vaccines may provide less effective protection against it, but disease severity may be lower than other strains^2^. A mechanistic modelling study^3^ found that the presence of immune-escaping variants significantly increases vaccination coverage thresholds needed to control transmission.

Moreover, uncertainties exist regarding the immune response to SARS-CoV-2. The duration of immunity for SARS-CoV-2 acquired from natural infection or vaccination is being actively studied. Evidence suggests that more than 90% of individuals with previous infection showed durable immune memory up to 5 months after infection^4^. Another study found that although the magnitude of immune response is heterogeneous, the immunity to SARS-CoV-2 is maintained at five months after infection^5^. A recent study of Pfizer-BioNTech’s vaccine suggested that its vaccine effectiveness gradually declined over 6 months after being fully vaccinated^6^, which has led to extensive debate about the need for vaccine booster doses^7^.

Under these uncertainties, appropriate combinations of vaccination and NPIs are needed to control SARS-CoV-2 transmission. A number of modeling studies have found that the effects of vaccination and NPIs depend on several factors, including vaccine rollout speed, vaccine effectiveness, and fraction of population protected by natural infection^8^. A mathematical model in King County, WA, suggested that rapid vaccination and adaptive lockdown can minimize future waves with the emergence of highly contagious SARS-CoV-2 variants^9^. Another age-structured transmission model that is calibrated to Portugal explored several sequential NPI policies while vaccines are being rolled out^8^.

Agent-based simulation has been employed in studies^10-12^ to account for heterogeneous individual behaviors and contact networks. These studies observe the impact of NPIs while vaccines are being rolled out. Li et al.^10^ presented a large-scale agent-based simulation to study the effects of different vaccine effectiveness, vaccine willingness, vaccine supply speed, and NPI policies in the U.S. However, model calibration was not provided. Another agent-based simulation model^11^ focused on the joint impact of COVID-19 vaccination and NPIs on infections, hospitalizations, and mortality with calibration using data from North Carolina. However, variants and immunity loss were not considered in both models. Most models’ NPI mitigation policies were somewhat simplistic, such as “maintain or remove” social distancing or changing social distancing levels at a certain point in time. In reality, the degree of NPIs may change continuously, either adaptively to the current epidemic situation, or based on a timeline coordinated with a vaccination schedule.

As a case study for a large urban area, we simulate COVID-19 transmission in King County, WA (greater Seattle) where regional parameters are calibrated to local epidemiological data from January 15, 2020 through December 31, 2020. Our agent-based simulation study uses detailed synthetic population data and includes interactions between vaccination and specific NPIs while considering immunity loss and variants. Our research investigates the impact of vaccination and NPI policies on COVID-19 deaths for several scenarios over four years (until the end of 2024). The scenarios include disease severity of the Omicron variant, immunity duration, NPI policy, vaccine willingness, booster schedule, and vaccine effectiveness against the Omicron variant. We aim to identify scenarios that yield an annual mortality rate from COVID-19 by 2024 that is on par with the target annual mortality rate from influenza and pneumonia.

## 2. Method

### 2.1. Model Overview

Our agent-based model is based on the open-source A Framework for Reconstructing Epidemics Dynamics (FRED) model^13^. We modified the FRED model to simulate SARS-CoV-2 transmission in King County, WA where approximately 1.9 million individuals are included^12^. The NPIs that we modeled include social distancing, face mask use, school closure, home quarantine, testing and contact tracing. In this paper, our natural history of COVID-19 follows a susceptible-exposed-infected-recovered (SEIR) model framework, including vaccination, SARS-CoV-2 variants, immunity loss from natural-infection or vaccination, comorbidities, and deaths (Figure 1).

**Figure 1.**
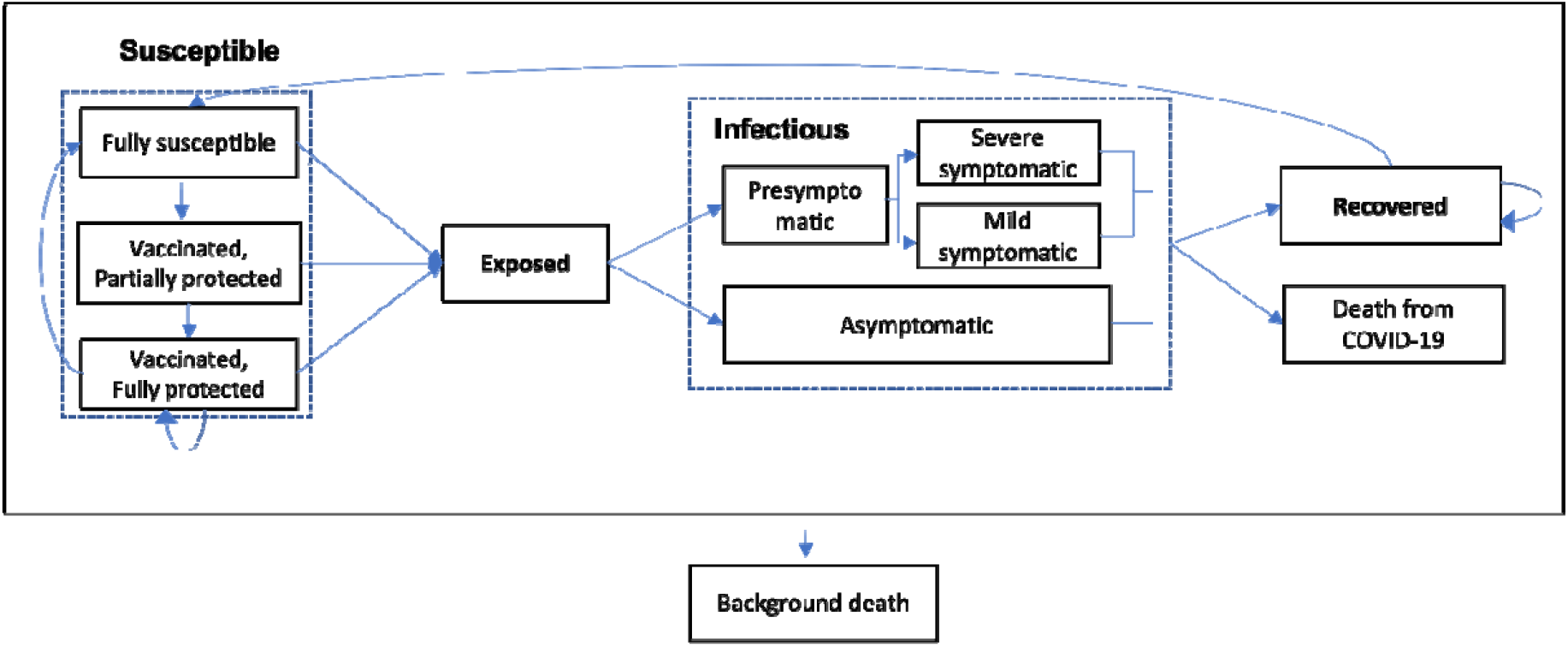
Natural history model of COVID-19 following a Susceptible-Exposed-Infected-Recovered (SEIR) framework.

#### Vaccination

We assume that anyone who is eligible and willing can be vaccinated unless the person is symptomatic on the day of vaccination. COVID-19 vaccines are known to be effective in preventing both infection and symptomatic disease, with greater effectiveness in preventing severe disease and death^14^. Thus, we model that susceptible individuals who are vaccinated develop immunity to SARS-CoV-2 and reduce the chance of symptomatic illness and death from SARS-CoV-2. Individuals with exposed or infectious status do not benefit from vaccination. When recovered individuals are vaccinated, they remain in the recovered state with immunity for a longer period. This accounts for ‘super immunity’ where individuals who are infected and vaccinated show extremely high levels of immune response^15^.

#### SARS-CoV-2 Variants

We consider three of the most widespread SARS-CoV-2 VOC, the Alpha, the Delta, and the Omicron variants. We model the Alpha variant as 29% more contagious and the Delta variant as 97% more contagious than the original strain^14,16^. Both variants were assumed to show the same disease severity as the original strain and the same vaccine effectiveness as the original strain^14^. The Omicron variant is assumed to be 60% more contagious than the Delta variant^14^. Disease severity and vaccines may exhibit reduced effectiveness against the Omicron variant.

Considering the impact of travelers who may introduce viruses from outside of King County, we import new infections based on a report from the Seattle-Tacoma International airport, ranging from 0 to 0.56 per day^17^. Based on Washington State’s SARS-CoV-2 sequencing results^18^, we assume that half of the new imports are VOC and the Alpha variant is introduced from January 1, 2021 to April 2, 2021, the Delta variant is introduced from April 3, 2021 to December 3, 2021, and the Omicron variant is introduced from December 4, 2021 to the end of the simulation.

#### Immunity loss

We model that immunity generated from vaccination or natural infection can be lost after some period. A gamma distribution is used to represent individuals’ immunity duration, to account for individuals’ heterogeneous immune responses^19,20^. In our model, when a person loses immunity, the person becomes fully susceptible. If a vaccinated individual is infected, the person loses vaccine-induced immunity but gains natural immunity once recovered.

#### Mortality

Deaths from COVID-19 and background mortality are considered. We assume that the infection-fatality ratio of COVID-19 depends on individuals’ age and comorbidity status to COVID-19^21,22^. Once an individual dies, the person is removed from each active location (household, neighborhood, school, and/or workplace) and no longer influences future transmission. Background death rate is based on gender and age^23^.

Values for all parameters and detailed transmission equations are given in the Appendix, Sections 1 and 2.

### 2.2. Calibration Procedure

We calibrate the model to data in the greater Seattle area from January 15, 2020, to December 31, 2020 by targeting basic reproduction number (R_0_) and reported deaths. We fit previous compliance history to NPIs by observing Seattle’s sequence of interventions. Parameters that we calibrate include COVID-19 transmissibility, contact rates at each location (household, neighborhood, school, and workplace), and default home quarantine percentage of symptomatic individuals. See Appendix 1.3 for the calibration procedure and detailed model description.

### 2.3. Parameter Settings for Scenario Analysis

Our simulation period spans five years, from January 15, 2020 (reported first day of infection in King County) to December 31, 2024. The period from January 15, 2020, to December 31, 2020, is calibrated. From January 1, 2021 to December 31, 2024 (four years), we perform a scenario analysis by simulating different vaccination and NPI policy parameters, with parameter settings listed in Table 1. For outcomes, we estimate daily median deaths and total deaths for the simulation period. In all analyses and calibration, we replicate 200 simulation runs with a different random seed.

**Table 1.** Parameter settings for scenario analyses

|  | **Parameter Settings** |
| --- | --- |
| Disease severity of the Omicron variant compared to other strains | Same, 50% less severe, 90% less severe |
| Average immunity duration | 9 months, 1 year |
| NPI policy | Timeline 1, Timeline 2, Threshold |
| Booster schedule | No booster, Booster every 6 months |
| Vaccine willingness | 60%, 80%, 100% |
| Vaccine effectiveness against the Omicron variant compared to other strains | Same, 25% less effective, 50% less effective |

#### Viral parameters

We model disease severity of the Omicron variant to be the same, 50% less, or 90% less compared to other strains^24^. If the disease is 90% less severe, an infected individual has a 90% reduced chance of developing symptoms, severe disease, and death. We model immunity duration from natural infection and vaccination using a gamma distribution. The shape and scale parameters are fitted as a mean value (shape times scale) of either 9 months or 1 year (Table 1)^24^. Furthermore, we fit the parameters to account for recent research^4^ showing that at least 90% of people have immunity to SARS-CoV-2 for 5 months after natural infection and vaccination.

#### NPI parameters

NPIs include social distancing, face mask use, school closure, home quarantine, testing and contact tracing. The effectiveness of face mask use is 25%. Although some studies show higher mask use effectiveness^25^, around 50 to 75%, we assumed that the effect of actual mask use is lower because people’s mask wearing behavior in a natural environment is not as strict as that of the experimental environment. We simulate different levels of compliance to social distancing, school closure, and face mask use by aggregating the three factors under one concept—NPI stage (Figure *2*). As the right panel of Figure *2* shows, we define four NPI stages ranging from a fairly closed society (stage 1) to a fully open society (stage 4). The left panel of Figure *2* shows NPI policies in which their timing of NPI stages differ. Timeline 1 gradually opens from 2021 to a fully open the society in 2022, whereas Timeline 2 opens more slowly than Timeline 1 and maintains some level of NPIs from 2022. We also introduce a Threshold NPI policy in which the NPI stage is dynamically determined by the last 2 weeks’ diagnosed infection cases. Under such a policy, the society is in stage 1 if diagnosed cases are higher than 350 cases per 100K population for 14-day rolling period, in stage 2 if there are 200 to 349 cases, in stage 3 if there are 100 to 199 cases, and in stage 4 if there are fewer than 100 cases. We fix compliance with home quarantine, testing, and contact tracing at the same level as those at the end of 2020.

**Figure 2.**
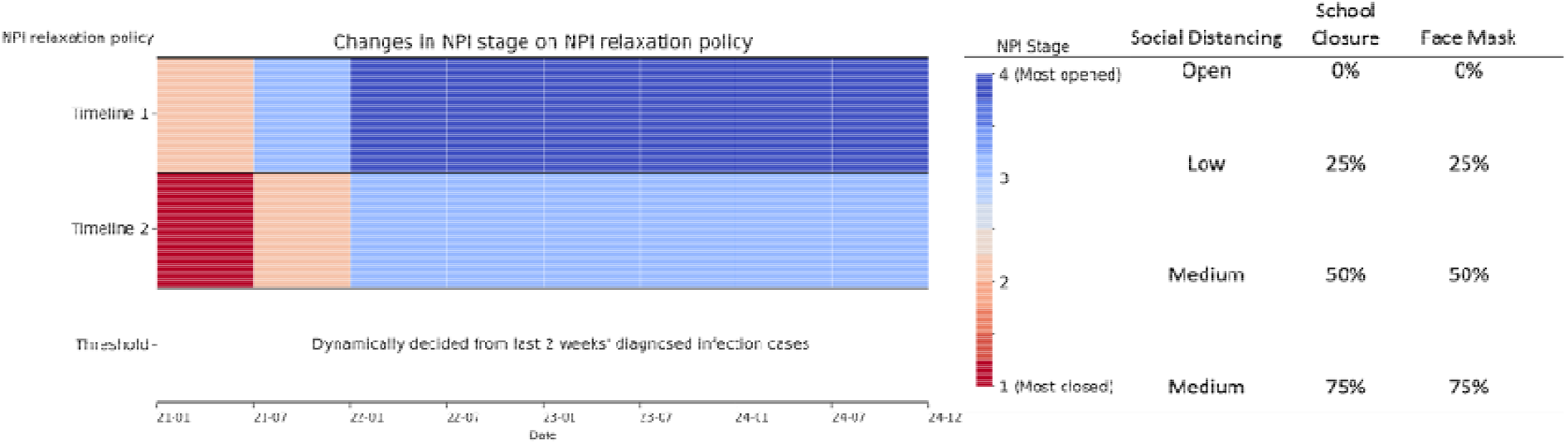
Definitions of NPI stages and timeline NPI policies. Date is in year-month.

#### Vaccination parameters

We model booster schedule in two cases; no boosters are scheduled after the second dose, and boosters are scheduled every 6 months after the second dose. Vaccine willingness is simulated at 60%, 80%, or 100%. In addition, we simulate different vaccine effectiveness against the Omicron variant with potentially lower vaccine effectiveness than other strains (original, Alpha, and Delta variants). Other settings are primarily based on the Pfizer-BioNTech COVID-19 vaccine. We assume that the vaccine has similar effects to other COVID-19 vaccines authorized by FDA for emergency use (e.g., Moderna). Thus, we assume that two doses are needed with the second dose administered 21 days after the first dose. Vaccine effectiveness is 51.4% on preventing infection and 54.4% on symptomatic diseases and deaths for the first dose after 12 days of vaccination^14,26^. For the second dose, the effectiveness increases to 86% on infection and 92% on symptomatic diseases and deaths after 7 days of vaccination^14^. When a booster dose is administered, we assume that it has the same level of vaccine effectiveness and immunity duration as the second dose. We followed Washington State’s vaccination schedule for prioritization policy and age-specific eligible date. See Appendix 1.2 for details.

We explore the impact of viral, NPI, and vaccination parameters on deaths during year 2021 to 2024. All parameter combinations from Table 1 yield 324 scenarios. To observe the endgame of the COVID-19 pandemic, our objective is to find scenarios where the mortality rate from COVID-19 is at or below the mortality rate of influenza and pneumonia. Because most of our scenarios show stable waves from 2023, we calculate annual mortality rate per 100K population between 2023 to 2024 for each scenario. Using the 2017 annual mortality rate of influenza and pneumonia of 12.6 per 100K population in Washington State^27^, we seek scenarios with annual mortality rate per 100K population during 2023 and 2024 below that target.

## 3. Result

Figure 3 presents the main results on simulated total deaths, assuming the average immunity duration is 9 months and a booster schedule of every 6 months. The complete results assuming 9 month and 1 year average immunity duration, with and without a booster plan, are available in the Appendix, Sections 3-6. The nine graphs in Figure 3 are organized into three columns for vaccine effectiveness (50% less, 25% less, and same effectiveness against the Omicron variant compared to other strains) and three rows for disease severity of Omicron compared to other strains (same, 50% less severe, and 90% less severe). Each of the nine graphs has three markers (squares, circles, and triangles) for vaccine willingness (60%, 80%, and 100%). The three NPI policies are plotted on the horizontal axis, and total deaths is plotted on the vertical axis.

**Figure 3.**
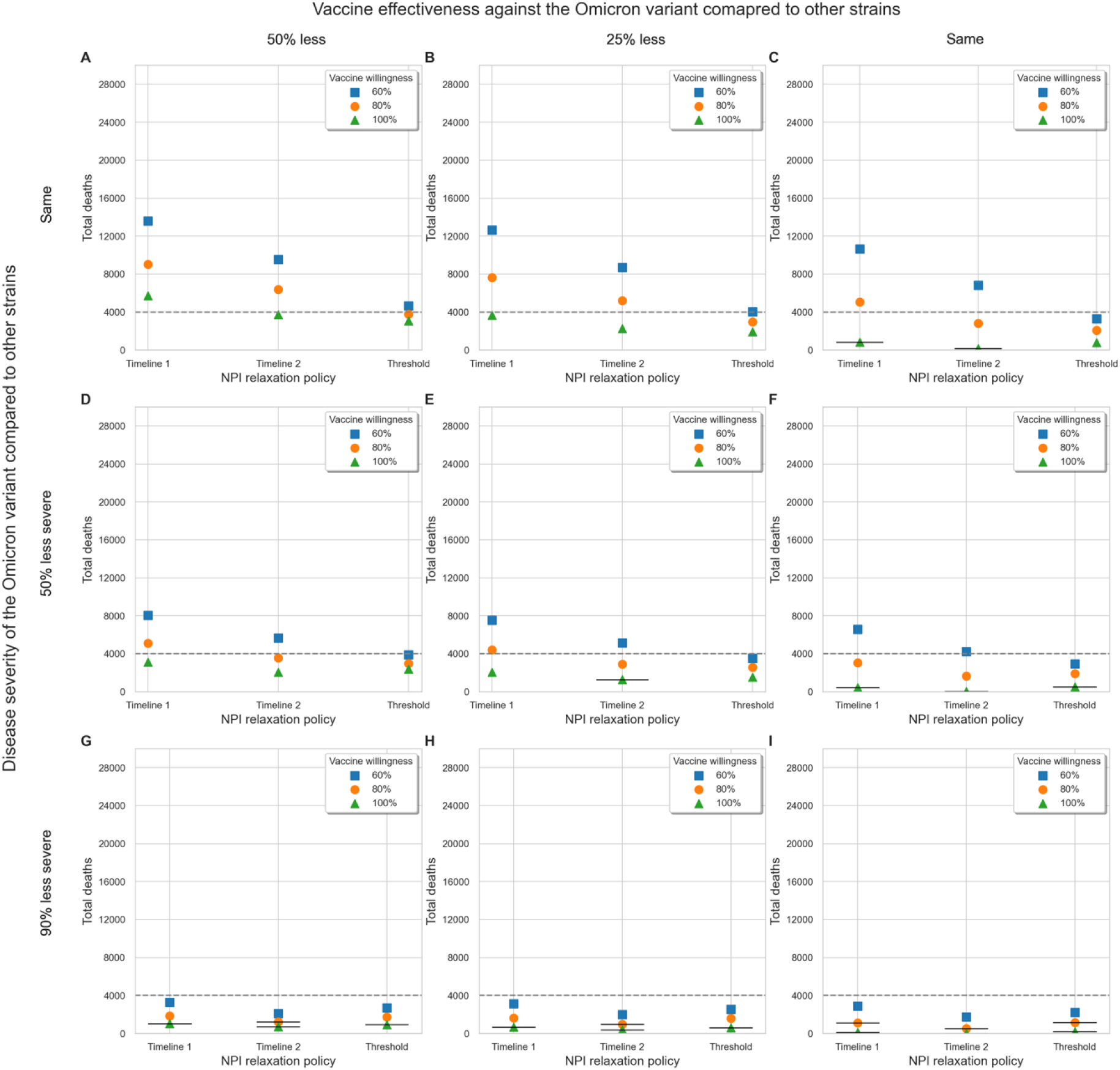
Impact of vaccine willingness and NPI policies on deaths from January 1, 2021, to December 31, 2024, with varying disease severity and vaccine effectiveness against the Omicron variant compared to other strains. Average immunity duration is assumed to be 9 months and boosters are scheduled every 6 months. Points (squares, circles, and triangles) marked with a short black line indicate scenarios where the annual mortality rate from 2023 to 2024 is below the target annual mortality rate from influenza and pneumonia in Washington State. To make the comparison easier, a total of 4,000 deaths (or 1,000 annual deaths) is grey lined.

### 3.1. Impact of viral characteristics

Figure 3 shows that the disease severity of the Omicron variant highly impacts death toll. Deaths are reduced by 64.9% on average when the variant is 50% less severe than having the same severity as other strains. If the variant is 90% less severe, the total deaths are further reduced by 131.5% compared to the variant having 50% less severity than other strains. If the average immunity duration is 1 year instead of 9 months, total deaths reduce by 35.8% on average. See Appendix, Section 5 and 6, for the results when the average immunity duration is 1 year and with or without boosters.

### 3.2. Impact of NPI policies

Compared to the Timeline 1 NPI policy which fully opens the society from January 2022, the Timeline 2 policy maintains NPI stage 3 from January 2022 to the end of 2024 (Figure *2*). With low-level of constant NPIs, the average total death toll is reduced by 35.1% compared to Timeline 1. The Threshold policy, which dynamically decides on the NPI stage according to real-time epidemic trend, further reduces the death toll by 62.8% compared to Timeline 2. This is due to the policy adaptively changing NPIs stages to avoid a resurgence. For example, in Figure 4, where the vaccine is 25% less effective against the Omicron variant and vaccine willingness is 80%, the result shows that Timelines 1 and 2 experience high peaks during early 2022 because the Omicron variant is rampant in the society but NPIs are relaxed regardless. However, the Threshold policy controls the epidemic by closing society when the daily cases are high.

**Figure 4.**
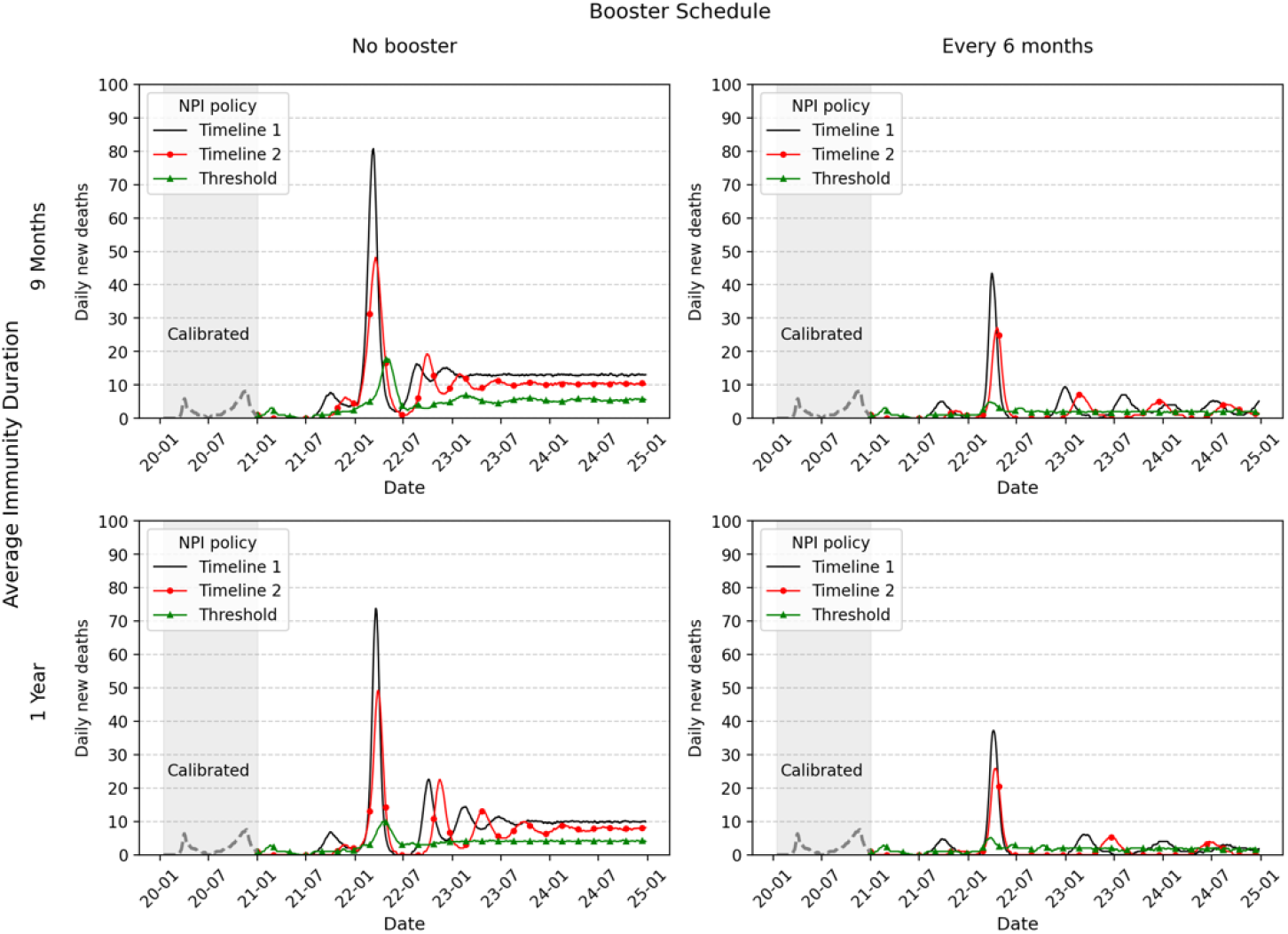
The impact of booster schedule, average immunity duration, and NPI policies on daily deaths. Vaccine is assumed to be 25% less effective against the Omicron variant than other strains and vaccine willingness is 80%.

Figure 5 illustrates the NPI stages in 2021-2024 for the Threshold policy that corresponds to the settings in Figure 3. The NPI stages shown in Figure 5 are averaged by 6-month intervals, where the red shading indicates NPI stage 1, or lockdown, and the blue shading indicates NPI stage 4, or total opening. For example, the Threshold policy in Figure 5A corresponds to the setting in Figure 3A, and the society is in NPI stage 1 and 2 when vaccine willingness is 60%. When the disease severity is 90% less (Figure 5G-5I), the society can be totally open from the second half of 2022 regardless of vaccine effectiveness and vaccine willingness. In general, the society can enter NPI stage 3 and 4 when the vaccine willingness is 100% regardless of vaccine effectiveness and disease severity of the Omicron variant.

**Figure 5.**
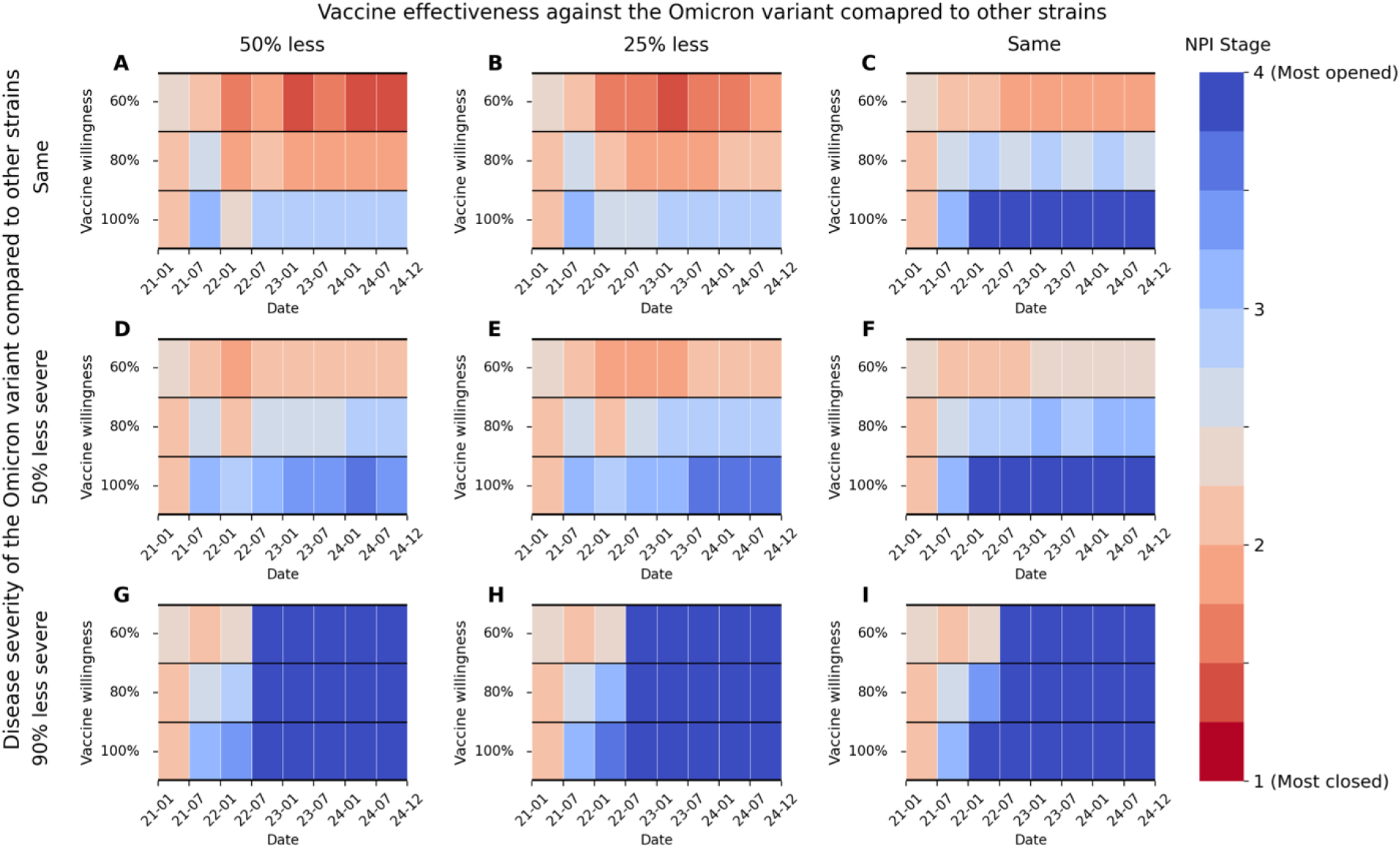
Changes in NPI stages (averaged by 6-month intervals) when Threshold NPI policy is applied with varying disease severity and vaccine effectiveness against the Omicron variant. compared to other strains. Average immunity duration is assumed to be 9 months and boosters are scheduled every 6 months (same setting as Figure 3).

Appendix, section 3.2 and 5.2, shows that when booster vaccines are not planned and Omicron has same or 50% less severity as other strains, the society is locked down to NPI stage 1 before 2024. Even if the Omicron variant’s disease severity is 90% less than other strains, NPI stage 4 (fully open) is not observed. When booster vaccines are planned every 6 months and the average immunity duration is 1 year, the NPI stages (shown in Appendix, Section 6.2) shows similar results to Figure 5 but are further relaxed.

### 3.3. Impact of booster schedule, vaccine willingness, and vaccine effectiveness against the Omicron variant compared to other strains

Among vaccine-related parameters, booster schedule has the greatest impact on mortality reduction. On average, total deaths are reduced by 267.7% when people receive booster vaccines every 6 months compared to not getting the booster. Figure 4 shows the effects of booster vaccine. With a booster schedule (right column of Figure 4), deaths are reduced from 2022. The effects of increasing vaccine willingness and vaccine effectiveness against the Omicron are magnified with the booster plan. If vaccine willingness increases from 60% to 80%, the average mortality decreases by 72.4%. The average mortality decreases by 118.4% if vaccine willingness increases from 80% to 100%. The increased vaccine willingness is more powerful when the Omicron variant is less severe, immunity duration is longer, vaccine has high effectiveness against the Omicron variant, and/or NPI policy is more relaxed. With the booster plan, the average reduction of total deaths is 19.9% when vaccine effectiveness against the Omicron variant is 25% less instead of 50% less than other strains. When the vaccine effectiveness increases from 25% less to same effectiveness as the other strains, the average reduction is 49.8%.

### 3.4. Can we end the pandemic?

Out of 324 scenarios, 52 scenarios yield a lower annual mortality rate from COVID-19 during 2023 to 2024 than the target goal of annual deaths from influenza and pneumonia in Washington State. Every satisfactory scenario has a 6-month booster schedule. Of the 52 scenarios, only 22 scenarios fully open the society from January 1, 2022, according to Timeline 1 or the Threshold NPI policy (Table 2). The 22 scenarios include combinations of Timeline 1 or Threshold NPI policy with vaccine willingness of 80% or above, low disease severity, long immunity duration, and high vaccine effectiveness against the Omicron variant. Only one scenario satisfies the objective when vaccine willingness is 60%. That scenario is when the NPI timeline is 2, boosters are scheduled every 6 months, the Omicron is 90% less severe than other strains, average immunity duration is 1 year, and the vaccine has the same effect on the Omicron variant as other strains (see Appendix Section 6.3).

**Table 2.** Scenarios that fully open the society from January 1, 2022 and yield lower annual mortality rate from COVID-19 during 2023 and 2024 than that from influenza and pneumonia in Washington State.

| Booster schedule | NPI policy | Vaccine willingness | Average immunity duration | Disease severity of the Omicron variant compared to other strains | Vaccine effectiveness against the Omicron variant compared to other strains | Annual death rate per 100 K |
| --- | --- | --- | --- | --- | --- | --- |
| Every 6 months | Timeline 1 | 80% | 9 months | 90% less | Same | 12.3 |
|  |  |  | 1 year | 90% less | 50% less effective | 8.1 |
|  |  |  |  |  | 25% less effective | 8.3 |
|  |  |  |  |  | Same | 5.4 |
|  |  | 100% | 9 months | Same | Same | 12.3 |
|  |  |  |  | 50% less | Same | 4.8 |
|  |  |  |  | 90% less | 50% less effective | 6.4 |
|  |  |  |  |  | 25% less effective | 3.3 |
|  |  |  |  |  | Same | 0.0 |
|  |  |  | 1 year | Same | 25% less effective | 10.0 |
|  |  |  |  |  | Same | 1.7 |
|  |  |  |  | 50% less | 50% less effective | 8.1 |
|  |  |  |  |  | 25% less effective | 4.0 |
|  |  |  |  |  | Same | 0.0 |
|  |  |  |  | 90% less | 50% less effective | 0.2 |
|  |  |  |  |  | 25% less effective | 0.0 |
|  |  |  |  |  | Same | 0.0 |
|  | Threshold | 100% | 9 months | 50% less | Same | 6.7 |
|  |  |  |  | 90% less | Same | 0.0 |
|  |  |  | 1 year | Same | Same | 0.4 |
|  |  |  |  | 50% less | Same | 0.0 |
|  |  |  |  | 90% less | Same | 0.0 |

## 4. Discussion

In this study, we present an agent-based simulation model for SARS-CoV-2 transmission and evaluate the impact of vaccination and NPIs on deaths over four years in a large urban area, King County, WA. In order for deaths from COVID-19 to be no higher than that of influenza and pneumonia, a booster schedule is essential. If we want to completely open the society from 2022 and achieve the mortality objective, the following conditions should be met. When vaccine willingness is 80%, disease severity of the Omicron variant should be 90% less than other strains. When vaccine willingness is 100%, several combinations of disease severity of the Omicron variant, high vaccine effectiveness or long immunity duration satisfy the objective.

Our research contributes to a better understanding of the effects of SARS-CoV-2 vaccination and NPI policies while considering variants and immunity duration. Using an agent-based simulation, our model captures individual heterogeneity in behaviors such as mask wearing and compliance with social distancing, risk factors such as age and comorbidity, and tracks viral variants. Rather than approximating the effects of NPIs as a single variable that changes the force of infection, a method commonly used in mathematical models for simplicity^8,9^, we separately model specific NPIs including social distancing, face mask use, school closure, testing, contact tracing, and home quarantine.

Our study highlights the crucial role of variants and immunity loss in understanding the impact of vaccination and NPI policies. As of January 2022, the highly transmissible Omicron variant is the most dominant strain in the U.S. and Washington State^18^. When its disease severity and vaccine effectiveness are unknown, booster vaccination schedule and careful NPI relaxation is important to reduce death toll. Without booster doses, a Threshold NPI policy implies that returning to lockdown is inevitable unless disease severity of the Omicron variant is 90% less than original strains. When a booster schedule is applied, on the other hand, deaths are reduced by approximately a third on average compared to without a booster. Under the 6-month booster schedule, the Threshold NPI policy rarely proceeds to lockdown.

Limitations exist in our model. We assume that recovered individuals have “all or no” immunity to SARS-CoV-2. After the immunity duration, a recovered person becomes fully susceptible. This approach has been tried in most modelling studies^20,28,29^ due to its simplicity and lack of knowledge about immune response over time. Recovered individuals may have different immune responses by strain type^2^. In addition, the immunity duration may depend on whether it was acquired by vaccination or natural infection. Considering that individuals who were previously infected and later vaccinated gain high immunity to SARS-CoV-2^15^, we extend the immunity duration of the recovered people if they are vaccinated in the recovered state. When the time course of the immune response is better understood, we could model immunity waning by time and strain type.

While we modeled the Alpha, the Delta, and the Omicron variant, other variants are emerging^2^. Our assumption that imported viruses have constant viral characteristics as the Omicron variant may not be true after 2022, and the number and subtypes of imported viruses may differ over time. If the disease prevalence decreases worldwide and the virus no longer enters the society, we could see an early end to the pandemic. However, if the pandemic continues with current trend, our study implies that we should be careful in treating COVID-19 as a seasonal influenza. If a variant that is more contagious than the Omicron variant is found, a proper combination of booster plan and NPI policies will be needed to gain herd immunity and protect the society.

Lastly, our model has simplified some individual behaviors in ways that may not reflect reality exactly. We modeled that vaccine willingness is the same across age and risk factors. In fact, willingness to vaccinate differs by age, underlying medical conditions, and race^30^. We also assumed that people’s vaccine willingness remains constant regardless of the number of doses, but willingness to get a booster vaccine appears to be less than the first two vaccinations. Individuals’ compliance with NPIs such as face mask use or social distancing were assumed to be independent of vaccination behavior which might not be true in reality^31,32^. Future research should consider heterogeneous human behavior for vaccination and NPIs.

In summary, our study provides estimated impacts of SARS-CoV-2 vaccination and NPI policies while considering variants and immunity loss using agent-based simulation. Although disease severity of the Omicron variant and immunity duration have a significant impact on the outbreak size, a booster vaccination schedule is needed to lower the mortality rate of COVID-19 at or below the mortality rate of influenza and pneumonia. If booster vaccines are not planned and the disease severity of the Omicron variant is not mild, some NPI policies will be needed to avoid large future outbreaks.

## Data Availability

All data produced in the present study are available upon reasonable request to the authors

## References

1. Thompson MG, Burgess JL, Naleway AL, et al. Interim estimates of vaccine effectiveness of BNT162b2 and mRNA-1273 COVID-19 vaccines in preventing SARS-CoV-2 infection among health care personnel, first responders, and other essential and frontline workers—eight US locations, December 2020–March 2021. Morbidity and Mortality Weekly Report. 2021;70(13):495.

2. Centers for Disease Control and Prevention. SARS-CoV-2 Variant Classifications and Definitions. 2021. [Online; Accessed 17-June-2021]. https://www.cdc.gov/coronavirus/2019-ncov/variants/variant-info.html#Concern

3. Cohen JA, Stuart RM, Nùñez RC, et al. Mechanistic modeling of SARS-CoV-2 immune memory, variants, and vaccines. medRxiv. 2021;doi:https://doi.org/10.1101/2021.05.31.21258018

4. Dan JM, Mateus J, Kato Y, et al. Immunological memory to SARS-CoV-2 assessed for up to 8 months after infection. Science. Feb 5 2021;371(6529) doi:10.1126/science.abf4063

5. Mazzoni A, Maggi L, Capone M, et al. Heterogeneous magnitude of immunological memory to SARS-CoV-2 in recovered individuals. Clin Transl Immunology. 2021;10(5):e1281. doi:10.1002/cti2.1281

6. Thomas SJ, Moreira ED, Kitchin N, et al. Six Month Safety and Efficacy of the BNT162b2 mRNA COVID-19 Vaccine. medRxiv. 2021:2021.07.28.21261159. doi:10.1101/2021.07.28.21261159

7. Food Drug Administration. FDA Authorizes Booster Dose of Pfizer-BioNTech COVID-19 Vaccine for Certain Populations. 2021. [Online; Accessed 17-Oct-2021]. https://www.fda.gov/news-events/press-announcements/fda-authorizes-booster-dose-pfizer-biontech-covid-19-vaccine-certain-populations

8. Viana J, van Dorp CH, Nunes A, et al. Controlling the pandemic during the SARS-CoV-2 vaccination rollout. Nat Commun. Jun 16 2021;12(1):3674. doi:10.1038/s41467-021-23938-8

9. Reeves DB, Bracis C, Swan DA, et al. Rapid vaccination and partial lockdown minimize 4th waves from emerging highly contagious SARS-CoV-2 variants. Med. 2021;2(5):573–574.

10. Li J, Giabbanelli P. Returning to a Normal Life via COVID-19 Vaccines in the United States: A Large-scale Agent-Based Simulation Study. JMIR Med Inform. Apr 29 2021;9(4):e27419. doi:10.2196/27419

11. Patel MD, Rosenstrom E, Ivy JS, et al. Association of Simulated COVID-19 Vaccination and Nonpharmaceutical Interventions With Infections, Hospitalizations, and Mortality. JAMA Network Open. 2021;4(6):e2110782–e2110782. doi:10.1001/jamanetworkopen.2021.10782

12. Lee S, Zabinsky ZB, Wasserheit JN, Kofsky SM, Liu S. COVID-19 Pandemic Response Simulation in a Large City: Impact of Nonpharmaceutical Interventions on Reopening Society. Med Decis Making. May 2021;41(4):419–429. doi:10.1177/0272989X211003081

13. Grefenstette JJ, Brown ST, Rosenfeld R, et al. FRED (a Framework for Reconstructing Epidemic Dynamics): an open-source software system for modeling infectious diseases and control strategies using census-based populations. BMC Public Health. Oct 8 2013;13:940. doi:10.1186/1471-2458-13-940

14. Institute for Health Metrics and Evaluation. COVID-19 model update: Omicron and waning immunity. 2021. [Online; Accessed 10-February-2022]. https://www.healthdata.org/special-analysis/omicron-and-waning-immunity

15. Bates TA, McBride SK, Winders B, et al. Antibody Response and Variant Cross-Neutralization After SARS-CoV-2 Breakthrough Infection. JAMA. 2022;327(2):179–181. doi:10.1001/jama.2021.22898

16. Campbell F, Archer B, Laurenson-Schafer H, et al. Increased transmissibility and global spread of SARS-CoV-2 variants of concern as at June 2021. Eurosurveillance. 2021;26(24):2100509.

17. Port of Seattle. Confirmed COVID-19 Positive Tests at SEA. https://www.portseattle.org/node/13520

18. Washington State, Department of Health. COVID-19 Variants. 2021. [Online; Accessed 5-September-2021]. https://www.doh.wa.gov/Emergencies/COVID19/Variants

19. Weitz JS, Beckett SJ, Coenen AR, et al. Modeling shield immunity to reduce COVID-19 epidemic spread. Nat Med. Jun 2020;26(6):849–854. doi:10.1038/s41591-020-0895-3

20. Blackwood JC, Cummings DA, Broutin H, Iamsirithaworn S, Rohani P. Deciphering the impacts of vaccination and immunity on pertussis epidemiology in Thailand. Proc Natl Acad Sci U S A. Jun 4 2013;110(23):9595–600. doi:10.1073/pnas.1220908110

21. Clark A, Jit M, Warren-Gash C, et al. How many are at increased risk of severe COVID-19 disease? Rapid global, regional and national estimates for 2020. MedRxiv. 2020;

22. A FAIR Health White Paper, Marty Makary. Risk Factors for COVID-19 Mortality among Privately Insured Patients: A Claims Data Analysis. FAIR Health, Inc 2020. [Online; Accessed 31-May-2021] https://s3.amazonaws.com/media2.fairhealth.org/whitepaper/asset/Risk%20Factors%20for%20COVID-19%20Mortality%20among%20Privately%20Insured%20Patients%20-%20A%20Claims%20Data%20Analysis%20-%20A%20FAIR%20Health%20White%20Paper.pdf

23. Flagg LA, Anderson RN. National Vital Statistics Reports. National Vital Statistics Reports 2021. [Online; Accessed 31-May 2021]. https://www.cdc.gov/nchs/data/nvsr/nvsr69/nvsr69-14-508.pdf

24. Centers for Disease Control and Prevention. COVID-19 Scenario Modeling Hub - Scenario Definitions 2022. [Online; Accessed 8-February-2022]. https://covid19scenariomodelinghub.org/viz.html

25. Prevention CfDCa. Science Brief: Community Use of Cloth Masks to Control the Spread of SARS-CoV-2. 2021. [Online; Accessed 22-October-2021]. https://www.cdc.gov/coronavirus/2019-ncov/science/science-briefs/masking-science-sars-cov2.html

26. Chodick G, Tene L, Patalon T, et al. Assessment of Effectiveness of 1 Dose of BNT162b2 Vaccine for SARS-CoV-2 Infection 13 to 24 Days After Immunization. JAMA Netw Open. Jun 1 2021;4(6):e2115985. doi:10.1001/jamanetworkopen.2021.15985

27. Centers for Disease Control and Prevention. Stats of the State of Washington. 2018. [Online; accessed 9-Feb-2022]. https://www.cdc.gov/nchs/pressroom/states/washington/washington.htm

28. Batistela CM, Correa DPF, Bueno AM, Piqueira JRC. SIRSi compartmental model for COVID-19 pandemic with immunity loss. Chaos Solitons Fractals. Jan 2021;142:110388. doi:10.1016/j.chaos.2020.110388

29. Kissler SM, Tedijanto C, Goldstein E, Grad YH, Lipsitch M. Projecting the transmission dynamics of SARS-CoV-2 through the postpandemic period. Science. 2020;368(6493):860–868.

30. Kelly BJ, Southwell BG, McCormack LA, et al. Predictors of willingness to get a COVID-19 vaccine in the U.S. BMC Infect Dis. Apr 12 2021;21(1):338. doi:10.1186/s12879-021-06023-9

31. Centers for Disease Control and Prevention. When You’ve Been Fully Vaccinated: How to Protect Yourself and Others. 2021. [Online; Accessed 22-June-2021]. https://www.cdc.gov/coronavirus/2019-ncov/vaccines/fully-vaccinated.html

32. Buckley J. Unlocking the World: EU Digital Covid Certificate: Everything you need to know. CNN; 2021. [Online; Accessed 22-June-2021]. https://www.cnn.com/travel/article/eu-covid-certificate-travel-explainer/index.html

