## Supplemental for "COVID-19 Endemic Plan: Impact of Vaccination and Non-pharmaceutical Interventions with Viral Variants and Waning Immunity Using an Agent-Based Simulation"

**Appendix**

### 1. COVID-19 Model Specification and Data Input

#### 1.1. Disease Parameters

Table 1 COVID-19 Disease Parameters

| **Disease Parameters** | **Value** | **Reference** |
| --- | --- | --- |
| Reproduction Number (R_0_) | 2 - 4 | ^1^ |
| Transmissibility of SARS-CoV-2 | 9.8% | Calibrated |
| Increased transmissibility of SARS-CoV-2 variant compared to original strain | Alpha variant - 29% higher  Delta variant - 97% higher  Omicron variant – 315% higher | ^2,3^ |
| Disease severity of SARS-CoV-2 variant compared to original strain | Alpha variant - Same  Delta variant - Same  Omicron variant – Varies (Same, 50% less, or 90% less) | ^4^ |
| Incubation period distribution | Lognormal with median of 5.1 days, dispersion of 1.52 days | ^5^ |
| Symptomatic period | 13 days | ^6^ |
| Pre-symptomatic period | 2 days | ^7^ |
| Infectious period | 15 days | ^7,8^ |
| Daily infectivity for 0-14 days | 0.5, 0.69, 0.94, 0.8, 0.61, 0.47, 0.35, 0.27, 0.2, 0.16, 0.12, 0.09, 0.07, 0.05, 0.04 | ^7-9^ |
| Infectiousness of asymptomatic individuals relative to symptomatic | 0.75 | ^1^ |
| Daily household contact probability | 85.6% | Calibrated |
| Daily contact rate in neighborhood | 1.188 | Calibrated |
| Daily contact rate in school^1^ | 3.508 | Calibrated |
| Daily contact rate in workplace^1^ | 1.745 | Calibrated |
| Probability of asymptomatic infection by age | 0-9: 0.71, 10-19: 0.79, 20-29: 0.73, 30-39: 0.67, 40-49: 0.6, 50-59: 0.51,60-69: 0.37, 70+: 0.31 | ^10^ |
| Probability of severe symptoms by age | 0-49: 0.017, 50-64: 0.045, 65+: 0.074 | ^11^ |
| Immunity duration (natural infection or vaccination) distribution when average immunity duration is 9 months and 1 year^2^ | Gamma distribution with shape and scale $\left( k,\theta\right)$  $\left( k,\theta\right)=\left( 7, 39.11 \right)$ for average of 9 months  $\left( k,\theta\right)=\left( 3.7, 98.65 \right)$ for average of 1 year | ^12^, Assumed |
| Days from infection to death^3^ | 15 days | ^11^ |
| Infection fatality ratio by age and comorbidity^4^ | Age \| Comorbidity to COVID-19 \| \|; \| None \| One \| Multiple; 0-14 \| 0.003% \| 0.01% \| 0.02%; 15-49 \| 0.02% \| 0.03% \| 0.09%; 50-54 \| 0.31% \| 0.54% \| 1.67%; 55-59 \| 0.28% \| 0.48% \| 1.49%; 60-64 \| 0.25% \| 0.42% \| 1.31%; 65-69 \| 0.21% \| 0.37% \| 1.14%; 70 + \| 1.83% \| 3.17% \| 9.77% | ^1,13,14^ |
| Background annual death rate by age and gender^5^ | Age \| Female \| Male \| Age \| Female \| Male; Under 1 year \| 0.5% \| 0.61% \| 45-54 \| 0.3% \| 0.49%; 1-4 \| 0.02% \| 0.03% \| 55-64 \| 0.67% \| 1.12%; 5-14 \| 0.01% \| 0.01% \| 65-74 \| 1.42% \| 2.2%; 15-24 \| 0.04% \| 0.10% \| 75-84 \| 3.79% \| 5.16%; 25-34 \| 0.08% \| 0.18% \| 85+ \| 12.87% \| 14.5%; 35-44 \| 0.14% \| 0.25% \| \| \| | ^15^ |

^1^ Workplace contacts include office contacts which is a smaller mixing network with twice of the workplace contact rate per day. Same policy is applied for school which includes classroom contact.

^2^ $k$ and $\theta$ were fitted to have mean value ($k\times\theta$) of 273 days, or 365 days, respectively, and at least 90% of immunity duration is more than 152 days (5 months)^12^.

^3^ Calculated from summing 12.9 days from symptom onset to death for 65 or more years plus 2 days of presymptomatic period^11^.

^4^ Based on the probability of death by age, $p\left( D | A \right)$^1^, probability of comorbidity (None, one, multiple) by age $p\left( C_{0,1,M} | A \right)$^13^, and odds ratios for infection fatality by comorbidity $p\left( C_{1,M}, D | C_{0}, D \right)$^14^. Given these values, $p\left( D | A \right)= p\left( C_{0} | A \right)\cdot p\left( D | C_{0},A \right)+p\left( C_{1} | A \right)\cdot p\left( D | C_{1},A \right)+p\left( C_{M} | A \right)\cdot p\left( D | C_{M},A \right)$. Using odds ratios for infection fatality by comorbidity, $p\left( D | C_{0},A \right)$ is derived by calculating $p\left( D | A \right)= p\left( C_{0} | A \right)\cdot p\left( D | C_{0},A \right)+p\left( C_{1} | A \right)\cdot p\left( D | C_{0},A \right)\cdot p\left( C_{1},D | C_{0},D \right)+p\left( C_{M} | A \right)\cdot p\left( D | C_{0},A \right)\cdot p\left( C_{M},D | C_{0},D \right)$. Then, infection fatality ratio of one or multiple comorbidities are calculated from $p\left( D | C_{1,M},A \right)= p\left( D | C_{0},A \right)\cdot p\left( C_{1,M}, D | C_{0}, D \right)$.

^5^ Converted annual death rate by age and gender (r_a,g_) to annual death probability ($1-exp(r_{a,g}))$ for each individual.

#### 1.2. Other Parameters

Table 2 Other parameters

| **Timeline Parameters** | **Value** | **Reference** |
| --- | --- | --- |
| Simulation Period | January 15, 2020, to December 31, 2024 | Assumed |
| Initial infector setting | 35-year old who was infectious from January 15, 2020, and was isolated 5 days later | ^16^ |
| Daily imported new infections^1^ | January to June 2020: 0  July to September 2020: 0.025  October 2020 to December 2024: 0.558 | ^17^ |
| Proportion new imports are SARS-CoV-2 variants of concern (VOC) from January 1, 2021 | 1/2 | Assumed |
| Imported VOC types | January 1, to April 2, 2021: Alpha  April 3, 2021 to December 3, 2021: Delta  December 4, 2021 to December 31, 2024: Omicron | Assumed, ^18^ |
| **NPI Parameters** | **Value** | **Reference** |
| Total diagnosed cases per 100K population in two weeks to enter phase 2,3,4 on Threshold NPI relaxation | Under 350, 200, 100 | ^19^ |
| Minimum policy duration on Threshold NPI relaxation | 3 weeks | Assumed |
| Effectiveness of wearing face mask^2^ | 25% | ^20^ |
| Home quarantine duration when contact traced | 14 days since last contact with a person who has COVID-19 | ^3^ |
| Home quarantine percentage when symptomatic without testing | 19.5% | Calibrated |
| Reduction in household contact when home quarantine from March 1, 2020 | 50% less | Assumed |
| Delay from onset to testing | 2 days | ^21^ |
| Delay to receive test results | 1 day | ^21^ |
| Test sensitivity | 0.9 | Assumed |
| Infector’s compliance with contact tracing | 0.9 | Assumed |
| Contact tracing effectiveness (Household, school, workplace, neighborhood member’s compliance  with home quarantine and testing when contact traced) | 0.9, 0.72, 0.16, 0.10 | ^22,23^ |
| Delays in reaching contacts | 2 days | ^21^ |
| **Vaccination Parameters** | **Value** | **Reference** |
| Vaccine prioritization policy | Region A’s phased priority (See below for details) | ^24,25^ |
| Number of vaccine dosage | No booster: 2 doses  Booster vaccines for every 6 month: 9 doses at maximum | ^26^ |
| Vaccine dose interval after previous dose | 21 days for 2^nd^ dose / 6 months for booster doses | ^26^ |
| Vaccine effect | Immune and disease-modifying | ^27^ |
| Vaccine effectiveness on preventing infection | 51.4% after 12 day of 1^st^ dose administration,  86% after 7 days of 2^nd^ or more dose administration | ^28^, Assumed for booster dose |
| Vaccine effectiveness on preventing symptomatic disease and death | 54.4% after 12 day of 1^st^ dose administration,  92% after 7 days of 2^nd^ or more dose administration | ^28^, Assumed for booster dose |
| Vaccine willingness of next dosage once received first dose | 100% | Assumed |
| Vaccine dosage policy | Additional dose recipients are prioritized over unvaccinated people on each day | Assumed |
| Vaccine supply schedule | See below | ^29^ |

^1^Port of Region A publishes the number of airline passengers who test positive for COVID-19 every three months from January 2020. Assuming detection rate is 40%^23^, we estimated daily infections by multiplying 2.5 (detection rate) and dividing by 90 days. Data on 2021 and 2022 were not available at the time of this study, so we fixed the values at the same level as that of October to December 2020.

^2^Assuming people wear surgical masks or cloth masks.

- Vaccine prioritization policy

We modeled vaccine prioritization policies based on Region A’s phased policy^25^. The phased priority order is as follows: 1) 65 years or older, or 50 years or older and household size is 4 or more, 2) 50 years or older or essential workers, 3) adults with two or more comorbidities, 4) essential workers younger than 50, 5) essential workers or have one comorbidity, and 6) everyone else.

- Vaccine eligibility date by age

Individuals are assumed to be eligible for the vaccine if they are ages 16 and older from January 1, 2021, ages 12 and over from May 12, 2021^30^, and ages 5 and over from November 1, 2021^31^. Children under 5 are currently under approval status. We assumed that vaccines for the population is available from July 1, 2022^32^.

- Vaccine supply schedule

Our daily dose of vaccines is based on the actual vaccine schedule administered by Region A from January to March 2021, after which the vaccine supply is the same as at the end of March 2021^29^. We adjusted the numbers considering that the synthetic population is based on 2010 census data. Figure 1 shows the simulated vaccine supply schedule.

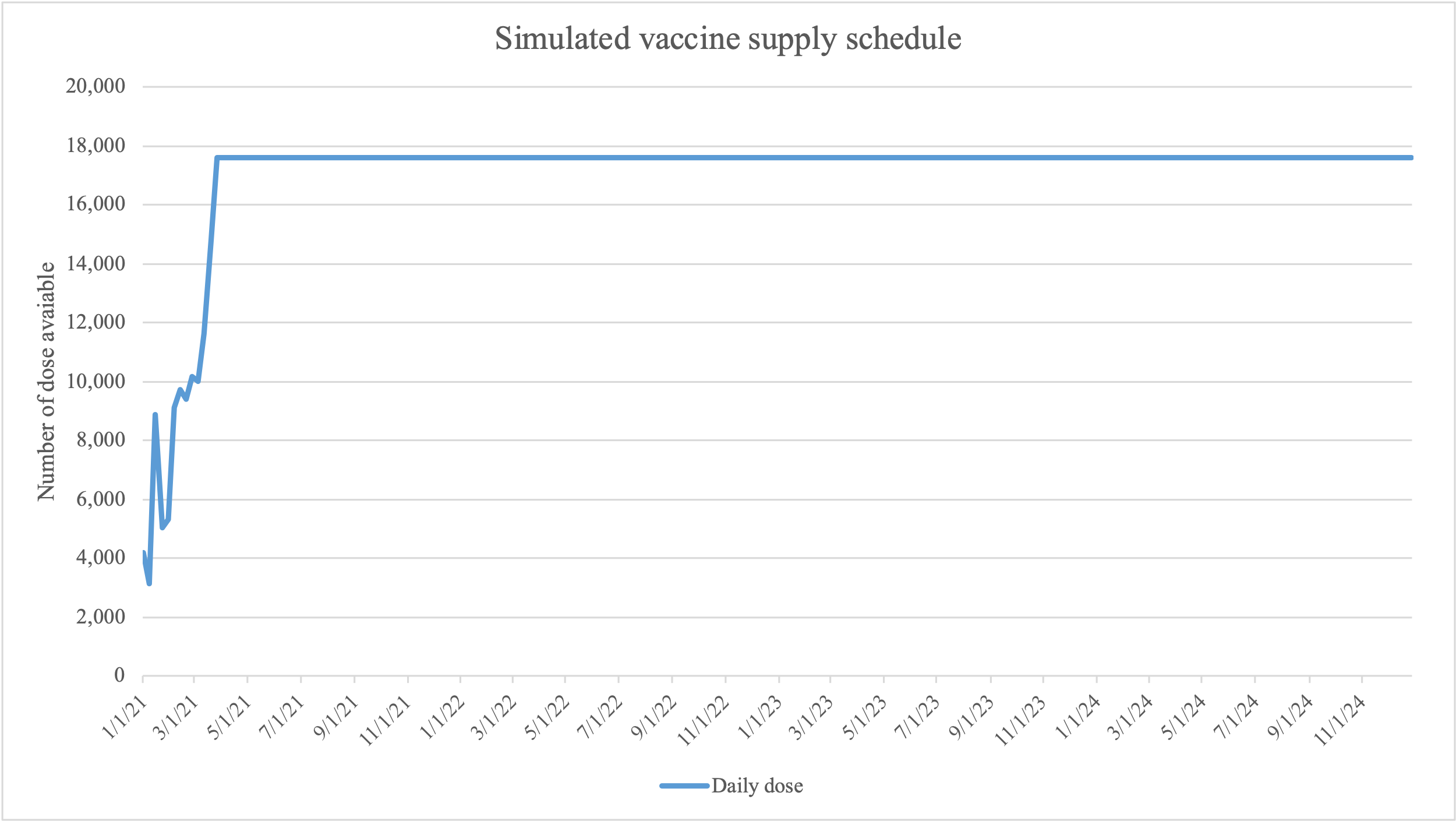

Figure 1 Simulated vaccine supply schedule

#### 1.3. Parameter Calibration

We calibrated the following unknown model parameters: transmissibility of SARS-CoV-2, daily contact probability between household members, daily contact rates of neighborhood, school, and workplace, and default home quarantine percentage of symptomatic individuals. In the first step, we sampled 1,000 candidate parameter sets using the Latin hypercube method and simulated each parameter set with 200 replications. We initially infected 10 random individuals and simulated for 30 days and identified average secondary infections caused by the initial infectors to calculate R_0_. Through this process, 55 parameter sets yielded R_0_ values between 2 and 4, a known range for COVID-19^1^. Then, we used Minitab 19.2020.1 (Minitab LLC) to identify clusters of the 55 parameter sets using complete-linkage method. This resulted in 8 clusters.

In the second step, we simulated public health interventions in Region A from January 15, 2020, through December 31, 2020, by observing Region A’s sequence of interventions (See Figure 2). We fit compliance to social distancing using IHME’s data on Region A’s social distancing^33^. We assumed that compliance to social distancing is low if changes in mobility is -20 to -30%, medium if the change is -30 to -40%, and high if the change is over -40%. From late August to early October 2020, there was a change in mobility between -16% and -19% in some days, but considering the small differences, the social distance was kept low. Compliance to face mask use was also based on IHME data^33^. We identified the first days that face mask usage exceeds 25%, 50%, and 75%, respectively. School closure was based on Region A timeline^34^. Testing and contact tracing were based on Lee, et al.^23^ Finally, we selected the cluster with the smallest difference between the time-weighted sum of 7-day moving average reported deaths during the calibration period.

Figure 3 shows calibration results on reported deaths, hospitalizations, and diagnosed cases. Although reported deaths in the second half of 2020 appear slightly later and higher than the model outcomes, the later peak is likely due to the fact that the time from infection to death continues to increase with advances in medical care, and higher mortality could occur due to the congestion in hospitals that cause excess deaths. As hospitalizations and cases show similar trends to the reported data, we conclude our calibration was appropriate.

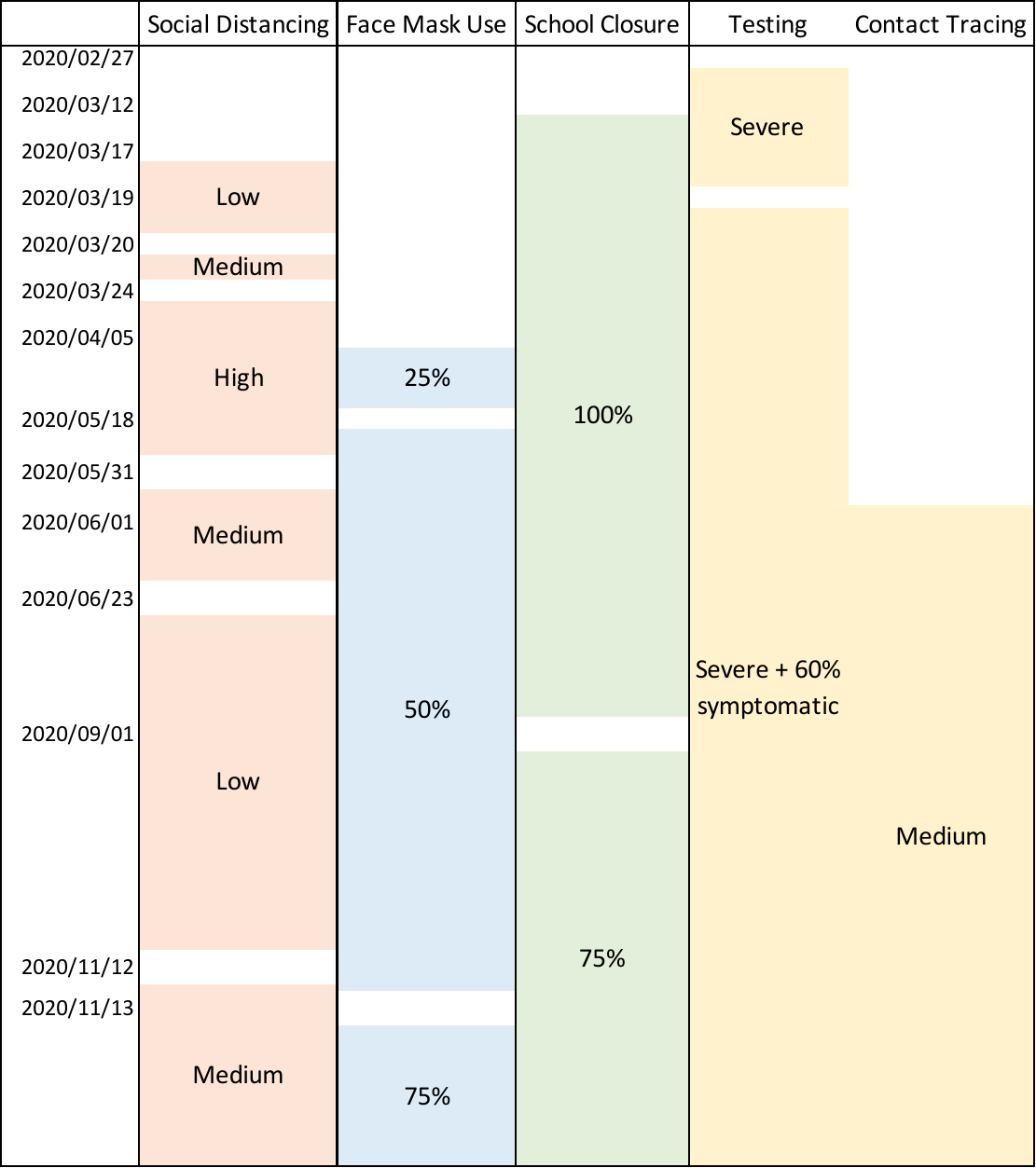

Figure 2 Diagram of simulated interventions from January 15, 2020, to December 31, 2020.

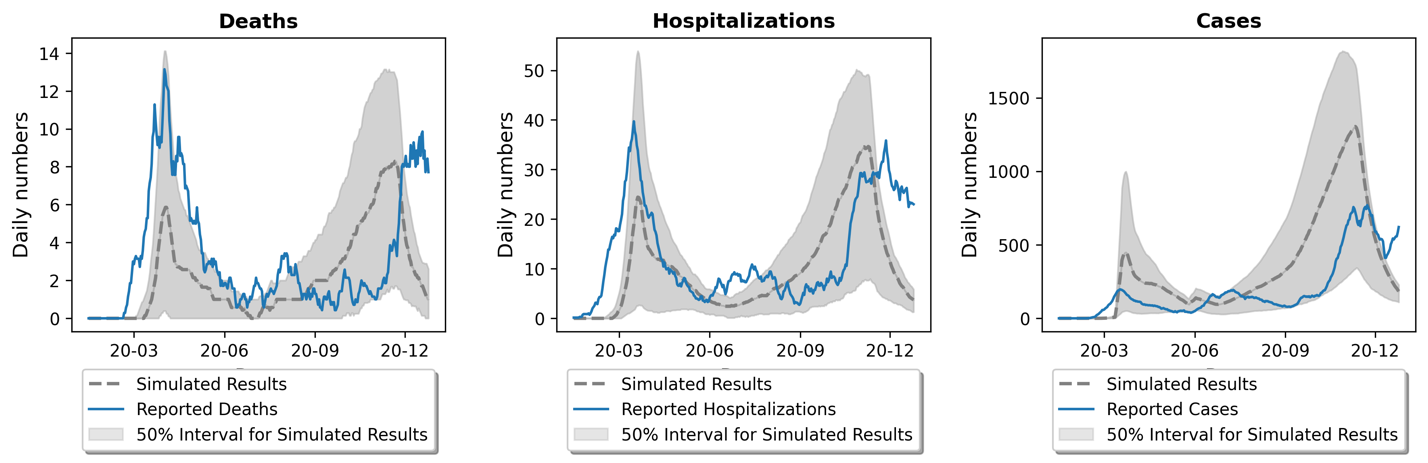

Figure 3 Calibration to reported deaths, hospitalizations, and cases in Region A. The 50% confidence interval was obtained by calculating 25% and 75% percentiles at each time step. Hospitalizations reported in Region A were based on the date of admission and model results were based on the date of infection. For comparison, we moved the reported hospitalizations to 12 days earlier, and graphed both outcomes based on the date of infection^1^.

### 2. SARS-CoV-2 Transmission Equations

- Household transmission

For an infectious person $i$, we consider his or her household transmission for each $d$th infectious day. For each susceptible household member $j$, $j$ is infected with $Household infection probability(i,j,d)$:

$$Household infection probability \left( i,j,d \right)=\left( Daily household contact probability \right)\times\left( 1-reduction in household contact if i is home quarantined \right) \times\left( transmissibility of SARS-CoV-2 \right)\times\left( Increased transmissibility of variant if i has variant \right)\times daily infectivity\left( i,d \right)\times\left( reduced infectiousness if i is asymptomatic \right) \times(1- vaccine effectiveness of preventing infection of j \times reduced vaccine effectiveness against variants if i has Omicron variant)$$

- Non-household transmission (Neighborhood, workplace, or school)

If an infectious person $i$ is present (not home quarantined) at the place $p$ on $d$th infectious day, the expected number of contacts is the $Contact count \left( i, p, d \right)$:

$$Contact count \left( i, p, d \right)=\left( Daily contacts per day \left( p \right) \right) \times\left( transmissibility of SARS-CoV-2 \right)\times\left( Increased transmissibility of variant if i has variant \right)\times daily infectivity\left( i,d \right)\times(reduced infectiousness if i is asymptomatic)$$

If $Contact count \left( i, p, d \right)$ is not integer, stochastic rounding is needed. If $Contact count \left( i, p, d \right)=x$,

$$Actual contact count (i,p,d)=Round(x)=\left\{ \begin{matrix} \lfloor x\rfloor\text{ with probability }1-(x-\lfloor x\rfloor) \\ \lfloor x\rfloor+1\text{ with probability }(x-\lfloor x\rfloor) \end{matrix} \right.$$

For example, if $x$ = 2.7, $Actual contact count (i,p,d)$ would be 2 with probability 0.3, and 3 with probability 0.7. We randomly select $Actual contact count (i,p,d)$ individuals in place $p$ on day $d$. For each contactee $j$ in the place $p$, if $j$ is susceptible and present in the place, $j$ is infected with $Non-household infection probability \left( i,j,d \right)$:

$$Non-household infection probability \left( i,j,d \right)=\left( 1-effectiveness of wearing face mask if i uses mask \right)\times(1-effectiveness of wearing face mask if j uses mask)\times(1-vaccine effectiveness of preventing infection of j \times reduced vaccine effectiveness against variants if i has Omicron variant)$$

### 3. Results – Average immunity duration is 9 months and no booster is scheduled

#### 3.1. Total Deaths

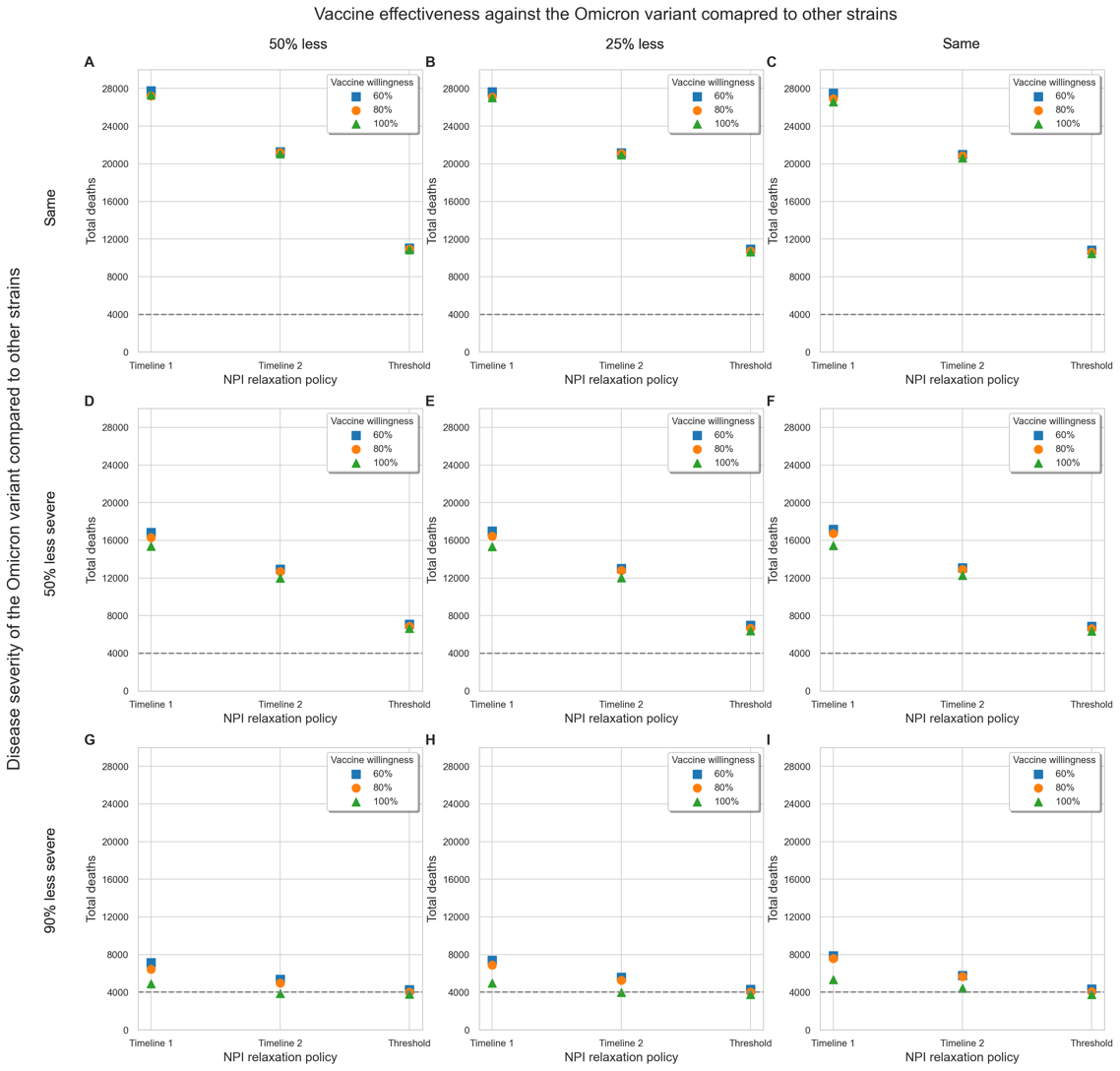

Figure 4 Impact of vaccine willingness and NPI policies on deaths from January 1, 2021, to December 31, 2024, with varying disease severity and vaccine effectiveness against the Omicron variant compared to other strains. Average immunity duration is assumed to be 9 months and no boosters are scheduled. Points (squares, circles, and triangles) marked with a short black line indicate scenarios where the annual mortality rate from 2023 to 2024 is below the target annual mortality rate from influenza and pneumonia in Region A. To make the comparison easier, a total of 4,000 deaths (or 1,000 annual deaths) is grey lined.

#### 3.2. Changes in NPI stages when Threshold NPI relaxation policy is applied

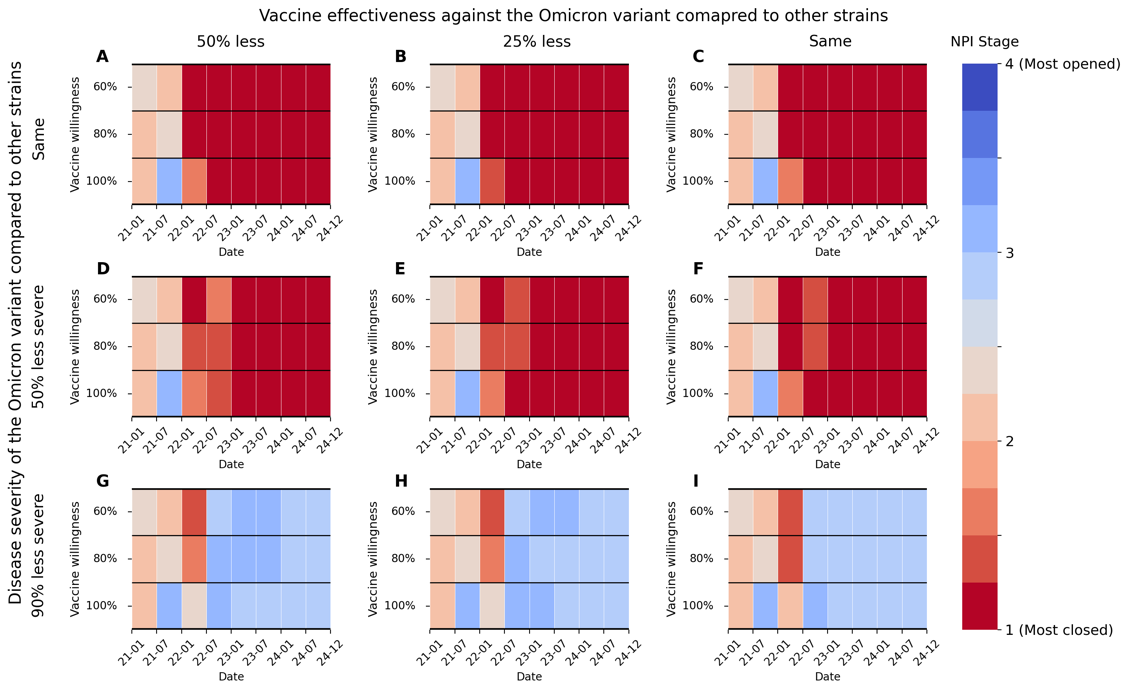

Figure 5 Changes in NPI stages (averaged by 6-month intervals) when Threshold NPI policy is applied with varying disease severity and vaccine effectiveness against the Omicron variant compared to other strains. Average immunity duration was assumed to be 9 months and booster is not scheduled.

#### 3.3. Detailed Results

Table 3 Detailed results when average immunity duration is 9 months and no booster is scheduled. Scenarios that meet the objective on reducing annual death rate from COVID-19 at or below the level of influenza and pneumonia (12.6) are colored red. Scenarios that fully open the society from January 1, 2022 are filled yellow.

| Disease severity of the Omicron variant compared to other strains | Vaccine effectiveness against the Omicron variant compared to other strains | NPI policy | Vaccine willingness | Deaths during 2021-2024 | Annual death rate per 100 K during 2023-2024 |
| --- | --- | --- | --- | --- | --- |
| Same | 50% less effective | 1 | 60 | 27,753 | 443.7 |
|  |  |  | 80 | 27,217 | 443.9 |
|  |  |  | 100 | 27,308 | 445.8 |
|  |  | 2 | 60 | 21,299 | 356.5 |
|  |  |  | 80 | 21,207 | 355.7 |
|  |  |  | 100 | 21,052 | 339.7 |
|  |  | Threshold | 60 | 11,036 | 186.1 |
|  |  |  | 80 | 10,936 | 185.3 |
|  |  |  | 100 | 10,858 | 183.2 |
|  | 25% less effective | 1 | 60 | 27,659 | 445.2 |
|  |  |  | 80 | 27,103 | 445.4 |
|  |  |  | 100 | 27,030 | 446.2 |
|  |  | 2 | 60 | 21,173 | 355.0 |
|  |  |  | 80 | 21,085 | 355.9 |
|  |  |  | 100 | 20,970 | 350.9 |
|  |  | Threshold | 60 | 10,958 | 188.8 |
|  |  |  | 80 | 10,739 | 187.0 |
|  |  |  | 100 | 10,650 | 185.1 |
|  | Same | 1 | 60 | 27,503 | 444.8 |
|  |  |  | 80 | 26,906 | 446.4 |
|  |  |  | 100 | 26,599 | 447.9 |
|  |  | 2 | 60 | 20,990 | 354.2 |
|  |  |  | 80 | 20,837 | 354.8 |
|  |  |  | 100 | 20,626 | 357.3 |
|  |  | Threshold | 60 | 10,851 | 187.8 |
|  |  |  | 80 | 10,624 | 187.2 |
|  |  |  | 100 | 10,464 | 185.9 |
| 50% less severe | 50% less effective | 1 | 60 | 16,854 | 249.1 |
|  |  |  | 80 | 16,290 | 249.7 |
|  |  |  | 100 | 15,366 | 251.0 |
|  |  | 2 | 60 | 12,948 | 200.3 |
|  |  |  | 80 | 12,698 | 200.5 |
|  |  |  | 100 | 11,970 | 191.5 |
|  |  | Threshold | 60 | 7,099 | 104.7 |
|  |  |  | 80 | 6,878 | 105.7 |
|  |  |  | 100 | 6,650 | 106.2 |
|  | 25% less effective | 1 | 60 | 16,983 | 249.7 |
|  |  |  | 80 | 16,456 | 250.1 |
|  |  |  | 100 | 15,352 | 251.8 |
|  |  | 2 | 60 | 13,018 | 199.6 |
|  |  |  | 80 | 12,799 | 200.9 |
|  |  |  | 100 | 12,011 | 197.3 |
|  |  | Threshold | 60 | 6,992 | 105.1 |
|  |  |  | 80 | 6,662 | 106.4 |
|  |  |  | 100 | 6,389 | 106.0 |
|  | Same | 1 | 60 | 17,168 | 249.1 |
|  |  |  | 80 | 16,716 | 250.1 |
|  |  |  | 100 | 15,434 | 252.2 |
|  |  | 2 | 60 | 13,091 | 198.8 |
|  |  |  | 80 | 12,921 | 198.4 |
|  |  |  | 100 | 12,278 | 201.7 |
|  |  | Threshold | 60 | 6,860 | 106.0 |
|  |  |  | 80 | 6,603 | 107.0 |
|  |  |  | 100 | 6,357 | 107.0 |
| 90% less severe | 50% less effective | 1 | 60 | 7,128 | 76.7 |
|  |  |  | 80 | 6,460 | 76.7 |
|  |  |  | 100 | 4,902 | 76.7 |
|  |  | 2 | 60 | 5,375 | 59.2 |
|  |  |  | 80 | 4,983 | 59.6 |
|  |  |  | 100 | 3,879 | 57.7 |
|  |  | Threshold | 60 | 4,276 | 58.2 |
|  |  |  | 80 | 4,012 | 58.4 |
|  |  |  | 100 | 3,795 | 58.4 |
|  | 25% less effective | 1 | 60 | 7,416 | 76.9 |
|  |  |  | 80 | 6,892 | 76.9 |
|  |  |  | 100 | 4,989 | 77.1 |
|  |  | 2 | 60 | 5,596 | 59.2 |
|  |  |  | 80 | 5,251 | 59.4 |
|  |  |  | 100 | 3,976 | 58.2 |
|  |  | Threshold | 60 | 4,310 | 58.0 |
|  |  |  | 80 | 4,031 | 58.2 |
|  |  |  | 100 | 3,774 | 58.2 |
|  | Same | 1 | 60 | 7,868 | 76.7 |
|  |  |  | 80 | 7,597 | 76.9 |
|  |  |  | 100 | 5,329 | 77.1 |
|  |  | 2 | 60 | 5,795 | 58.2 |
|  |  |  | 80 | 5,662 | 58.4 |
|  |  |  | 100 | 4,432 | 59.6 |
|  |  | Threshold | 60 | 4,344 | 57.8 |
|  |  |  | 80 | 4,056 | 57.8 |
|  |  |  | 100 | 3,755 | 58.0 |

### 4. Results – Average immunity duration is 9 months and booster is scheduled for every 6 months

#### 4.1. Total Deaths

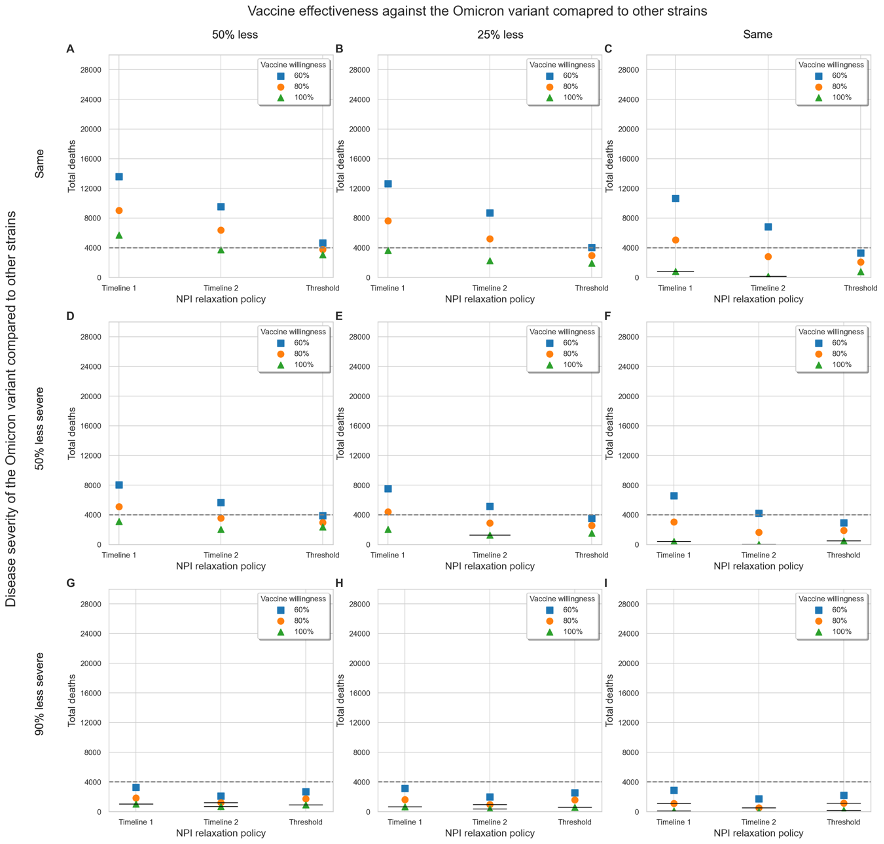

Figure 6 Impact of vaccine willingness and NPI policies on deaths from January 1, 2021, to December 31, 2024, with varying disease severity and vaccine effectiveness against the Omicron variant compared to other strains. Average immunity duration is assumed to be 9 months and boosters are scheduled every 6 months. Points (squares, circles, and triangles) marked with a short black line indicate scenarios where the annual mortality rate from 2023 to 2024 is below the target annual mortality rate from influenza and pneumonia in Region A. To make the comparison easier, a total of 4,000 deaths (or 1,000 annual deaths) is grey lined.

#### 4.2. Changes in NPI stages when Threshold NPI relaxation policy is applied

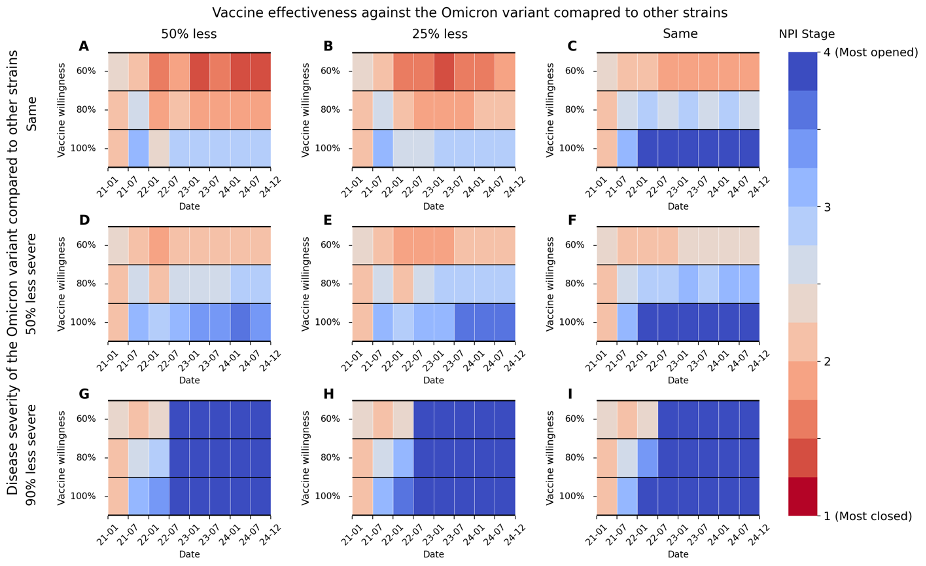

Figure 7 Changes in NPI stages (averaged by 6-month intervals) when Threshold NPI policy is applied with varying disease severity and vaccine effectiveness against the Omicron variant compared to other strains. Average immunity duration is 9 months and booster is scheduled for every 6 months.

#### 4.3. Detailed Results

Table 4 Detailed results when average immunity duration is 9 months and booster is scheduled for every 6 months. Scenarios that meet the objective on reducing annual death rate from COVID-19 at or below the level of influenza and pneumonia (12.6) are colored red. Scenarios that fully open the society from January 1, 2022 are filled yellow.

| Disease severity of the Omicron variant compared to other strains | Vaccine effectiveness against the Omicron variant compared to other strains | NPI policy | Vaccine willingness | Deaths during 2021-2024 | Annual Death Rate per 100 K during 2023-2024 |
| --- | --- | --- | --- | --- | --- |
| Same | 50% less effective | 1 | 60 | 13,609 | 190.5 |
|  |  |  | 80 | 9,015 | 109.5 |
|  |  |  | 100 | 5,719 | 60.0 |
|  |  | 2 | 60 | 9,541 | 128.2 |
|  |  |  | 80 | 6,388 | 86.6 |
|  |  |  | 100 | 3,739 | 32.6 |
|  |  | Threshold | 60 | 4,663 | 61.9 |
|  |  |  | 80 | 3,781 | 50.3 |
|  |  |  | 100 | 3,061 | 37.8 |
|  | 25% less effective | 1 | 60 | 12,654 | 181.1 |
|  |  |  | 80 | 7,619 | 104.3 |
|  |  |  | 100 | 3,675 | 38.2 |
|  |  | 2 | 60 | 8,697 | 127.1 |
|  |  |  | 80 | 5,194 | 77.5 |
|  |  |  | 100 | 2,276 | 22.4 |
|  |  | Threshold | 60 | 4,037 | 58.4 |
|  |  |  | 80 | 2,946 | 44.7 |
|  |  |  | 100 | 1,934 | 29.5 |
|  | Same | 1 | 60 | 10,662 | 165.6 |
|  |  |  | 80 | 5,045 | 86.6 |
|  |  |  | 100 | 808 | 12.3 |
|  |  | 2 | 60 | 6,832 | 116.3 |
|  |  |  | 80 | 2,801 | 47.0 |
|  |  |  | 100 | 159 | 3.3 |
|  |  | Threshold | 60 | 3,292 | 50.7 |
|  |  |  | 80 | 2,080 | 32.0 |
|  |  |  | 100 | 773 | 13.3 |
| 50% less severe | 50% less effective | 1 | 60 | 8,065 | 105.9 |
|  |  |  | 80 | 5,109 | 57.7 |
|  |  |  | 100 | 3,130 | 29.9 |
|  |  | 2 | 60 | 5,648 | 71.1 |
|  |  |  | 80 | 3,567 | 45.1 |
|  |  |  | 100 | 2,060 | 15.8 |
|  |  | Threshold | 60 | 3,899 | 52.8 |
|  |  |  | 80 | 2,969 | 38.6 |
|  |  |  | 100 | 2,369 | 27.4 |
|  | 25% less effective | 1 | 60 | 7,539 | 100.5 |
|  |  |  | 80 | 4,406 | 55.5 |
|  |  |  | 100 | 2,068 | 18.9 |
|  |  | 2 | 60 | 5,161 | 70.2 |
|  |  |  | 80 | 2,908 | 40.3 |
|  |  |  | 100 | 1,263 | 10.4 |
|  |  | Threshold | 60 | 3,509 | 50.9 |
|  |  |  | 80 | 2,582 | 37.2 |
|  |  |  | 100 | 1,521 | 21.0 |
|  | Same | 1 | 60 | 6,572 | 92.7 |
|  |  |  | 80 | 3,036 | 48.4 |
|  |  |  | 100 | 419 | 4.8 |
|  |  | 2 | 60 | 4,207 | 65.6 |
|  |  |  | 80 | 1,630 | 26.4 |
|  |  |  | 100 | 14 | 0.0 |
|  |  | Threshold | 60 | 2,924 | 42.8 |
|  |  |  | 80 | 1,910 | 27.6 |
|  |  |  | 100 | 500 | 6.7 |
| 90% less severe | 50% less effective | 1 | 60 | 3,294 | 32.0 |
|  |  |  | 80 | 1,829 | 15.6 |
|  |  |  | 100 | 1,019 | 6.4 |
|  |  | 2 | 60 | 2,107 | 19.7 |
|  |  |  | 80 | 1,211 | 12.1 |
|  |  |  | 100 | 699 | 4.0 |
|  |  | Threshold | 60 | 2,690 | 33.4 |
|  |  |  | 80 | 1,731 | 18.1 |
|  |  |  | 100 | 909 | 7.7 |
|  | 25% less effective | 1 | 60 | 3,113 | 30.5 |
|  |  |  | 80 | 1,615 | 15.2 |
|  |  |  | 100 | 676 | 3.3 |
|  |  | 2 | 60 | 1,993 | 20.6 |
|  |  |  | 80 | 974 | 10.2 |
|  |  |  | 100 | 385 | 1.3 |
|  |  | Threshold | 60 | 2,550 | 32.0 |
|  |  |  | 80 | 1,574 | 16.6 |
|  |  |  | 100 | 576 | 2.3 |
|  | Same | 1 | 60 | 2,862 | 29.1 |
|  |  |  | 80 | 1,109 | 12.3 |
|  |  |  | 100 | 124 | 0.0 |
|  |  | 2 | 60 | 1,720 | 19.9 |
|  |  |  | 80 | 505 | 5.6 |
|  |  |  | 100 | 11 | 0.0 |
|  |  | Threshold | 60 | 2,196 | 26.6 |
|  |  |  | 80 | 1,141 | 11.8 |
|  |  |  | 100 | 182 | 0.0 |

### 5. Results – Average immunity duration is 1 year and no booster is scheduled

#### 5.1. Total Deaths

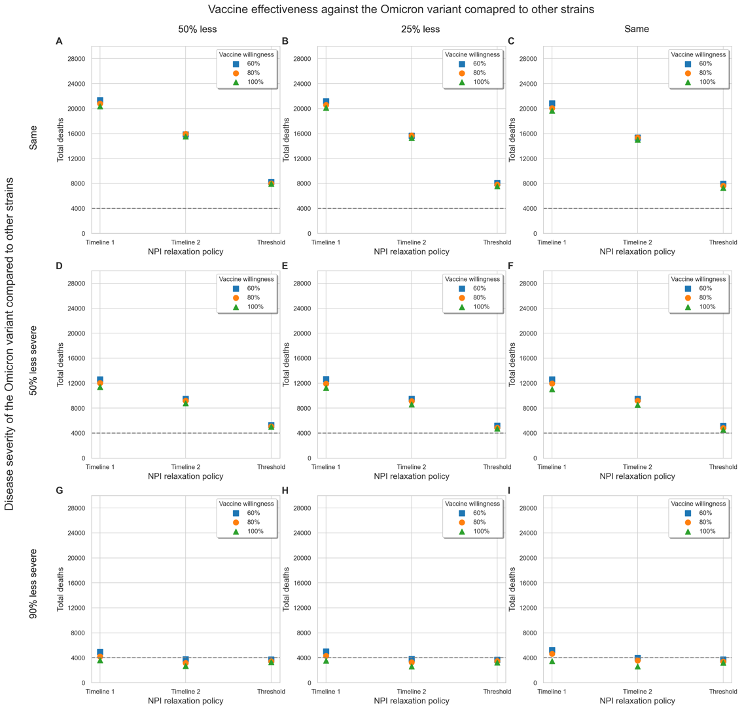

Figure 8 Impact of vaccine willingness and NPI policies on deaths from January 1, 2021, to December 31, 2024, with varying disease severity and vaccine effectiveness against the Omicron variant compared to other strains. Average immunity duration is assumed to be 1 year and no booster is scheduled..Points (squares, circles, and triangles) marked with a short black line indicate scenarios where the annual mortality rate from 2023 to 2024 is below the target annual mortality rate from influenza and pneumonia in Region A. To make the comparison easier, a total of 4,000 deaths (or 1,000 annual deaths) is grey lined.

#### 5.2. Changes in NPI stages when Threshold NPI relaxation policy is applied

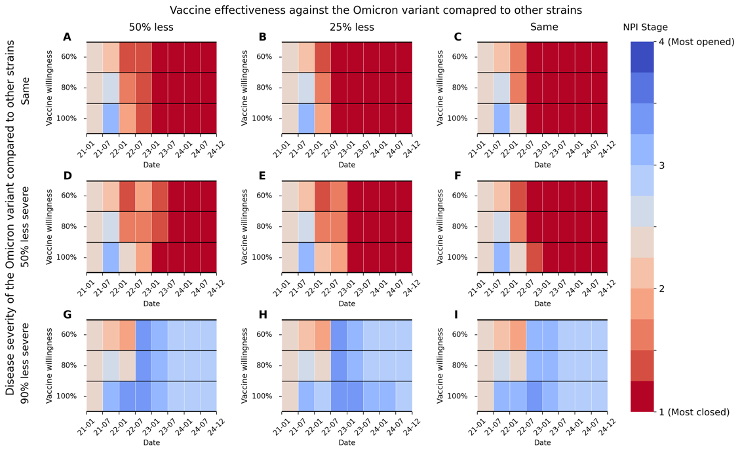

Figure 9 Changes in NPI stages (averaged by 6-month intervals) when Threshold NPI policy is applied with varying disease severity and vaccine effectiveness against the Omicron variant compared to other strains. Average immunity duration was assumed to be 1 year and booster is not scheduled.

#### 5.3. Detailed Results

Table 5 Detailed results when average immunity duration is 1 year and no booster is scheduled. Scenarios that meet the objective on reducing annual death rate from COVID-19 at or below the level of influenza and pneumonia (12.6) are colored red. Scenarios that fully open the society from January 1, 2022 are filled yellow.

| Disease severity of the Omicron variant compared to other strains | Vaccine effectiveness against the Omicron variant compared to other strains | NPI policy | Vaccine willingness | Deaths during 2021-2024 | Annual Death Rate per 100 K during 2023-2024 |
| --- | --- | --- | --- | --- | --- |
| Same | 50% less effective | 1 | 60 | 21,364 | 343.6 |
|  |  |  | 80 | 20,813 | 341.5 |
|  |  |  | 100 | 20,351 | 332.6 |
|  |  | 2 | 60 | 15,889 | 260.5 |
|  |  |  | 80 | 15,947 | 251.6 |
|  |  |  | 100 | 15,561 | 263.0 |
|  |  | Threshold | 60 | 8,255 | 138.2 |
|  |  |  | 80 | 7,994 | 139.0 |
|  |  |  | 100 | 7,972 | 138.2 |
|  | 25% less effective | 1 | 60 | 21,155 | 343.0 |
|  |  |  | 80 | 20,578 | 342.8 |
|  |  |  | 100 | 20,137 | 338.0 |
|  |  | 2 | 60 | 15,664 | 262.8 |
|  |  |  | 80 | 15,693 | 259.0 |
|  |  |  | 100 | 15,336 | 255.7 |
|  |  | Threshold | 60 | 8,091 | 137.1 |
|  |  |  | 80 | 7,823 | 138.1 |
|  |  |  | 100 | 7,551 | 138.6 |
|  | Same | 1 | 60 | 20,853 | 341.1 |
|  |  |  | 80 | 20,074 | 341.7 |
|  |  |  | 100 | 19,669 | 340.3 |
|  |  | 2 | 60 | 15,374 | 266.9 |
|  |  |  | 80 | 15,341 | 266.9 |
|  |  |  | 100 | 15,038 | 262.0 |
|  |  | Threshold | 60 | 7,954 | 136.5 |
|  |  |  | 80 | 7,627 | 138.4 |
|  |  |  | 100 | 7,307 | 138.2 |
| 50% less severe | 50% less effective | 1 | 60 | 12,628 | 191.9 |
|  |  |  | 80 | 12,027 | 191.1 |
|  |  |  | 100 | 11,407 | 186.1 |
|  |  | 2 | 60 | 9,490 | 144.8 |
|  |  |  | 80 | 9,200 | 140.8 |
|  |  |  | 100 | 8,794 | 144.2 |
|  |  | Threshold | 60 | 5,307 | 78.1 |
|  |  |  | 80 | 5,086 | 77.3 |
|  |  |  | 100 | 5,041 | 78.1 |
|  | 25% less effective | 1 | 60 | 12,645 | 191.9 |
|  |  |  | 80 | 11,926 | 191.9 |
|  |  |  | 100 | 11,255 | 188.2 |
|  |  | 2 | 60 | 9,480 | 146.5 |
|  |  |  | 80 | 9,117 | 143.3 |
|  |  |  | 100 | 8,608 | 142.9 |
|  |  | Threshold | 60 | 5,191 | 79.1 |
|  |  |  | 80 | 4,857 | 78.7 |
|  |  |  | 100 | 4,738 | 79.4 |
|  | Same | 1 | 60 | 12,609 | 189.5 |
|  |  |  | 80 | 11,938 | 190.3 |
|  |  |  | 100 | 11,029 | 190.1 |
|  |  | 2 | 60 | 9,471 | 148.3 |
|  |  |  | 80 | 9,186 | 148.3 |
|  |  |  | 100 | 8,512 | 146.2 |
|  |  | Threshold | 60 | 5,126 | 78.9 |
|  |  |  | 80 | 4,818 | 79.4 |
|  |  |  | 100 | 4,532 | 79.8 |
| 90% less severe | 50% less effective | 1 | 60 | 4,937 | 57.8 |
|  |  |  | 80 | 4,216 | 57.5 |
|  |  |  | 100 | 3,603 | 56.5 |
|  |  | 2 | 60 | 3,756 | 42.2 |
|  |  |  | 80 | 3,154 | 40.3 |
|  |  |  | 100 | 2,699 | 41.5 |
|  |  | Threshold | 60 | 3,731 | 53.2 |
|  |  |  | 80 | 3,437 | 52.6 |
|  |  |  | 100 | 3,313 | 53.4 |
|  | 25% less effective | 1 | 60 | 5,016 | 57.7 |
|  |  |  | 80 | 4,320 | 58.0 |
|  |  |  | 100 | 3,525 | 56.5 |
|  |  | 2 | 60 | 3,809 | 42.2 |
|  |  |  | 80 | 3,266 | 41.8 |
|  |  |  | 100 | 2,617 | 41.3 |
|  |  | Threshold | 60 | 3,695 | 52.4 |
|  |  |  | 80 | 3,451 | 53.0 |
|  |  |  | 100 | 3,260 | 52.6 |
|  | Same | 1 | 60 | 5,246 | 57.7 |
|  |  |  | 80 | 4,653 | 57.7 |
|  |  |  | 100 | 3,454 | 57.5 |
|  |  | 2 | 60 | 3,985 | 42.0 |
|  |  |  | 80 | 3,567 | 42.0 |
|  |  |  | 100 | 2,617 | 42.2 |
|  |  | Threshold | 60 | 3,710 | 52.8 |
|  |  |  | 80 | 3,400 | 52.6 |
|  |  |  | 100 | 3,200 | 52.6 |

### 6. Results – Average immunity duration is 1 year and booster is scheduled for every 6 months

#### 6.1. Total Deaths

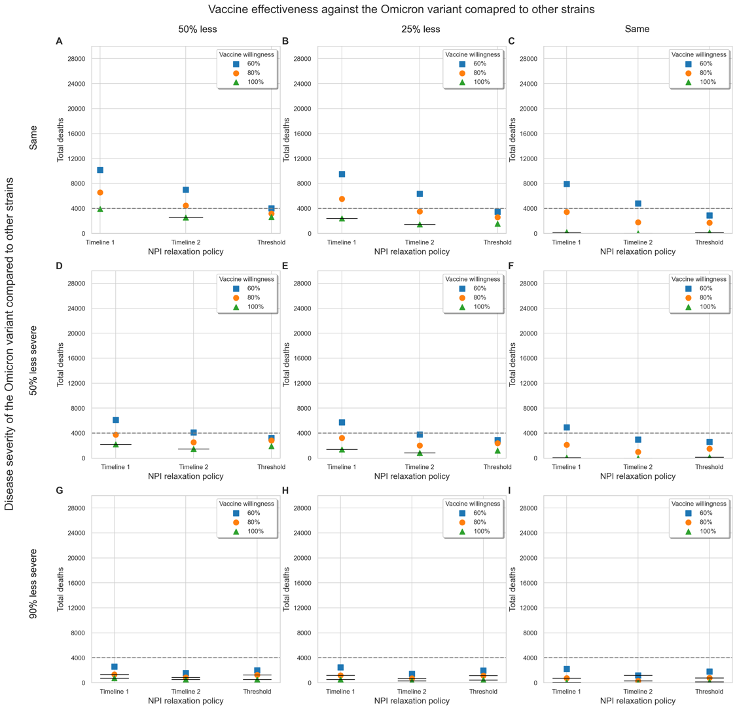

Figure 10 Impact of vaccine willingness and NPI policies on deaths from January 1, 2021, to December 31, 2024, with varying disease severity and vaccine effectiveness against the Omicron variant compared to other strains. Average immunity duration is assumed to be 9 months and boosters are scheduled every 6 months. Points (squares, circles, and triangles) marked with a short black line indicate scenarios where the annual mortality rate from 2023 to 2024 is below the target annual mortality rate from influenza and pneumonia in Region A. To make the comparison easier, a total of 4,000 deaths (or 1,000 annual deaths) is grey lined.

#### 6.2. Changes in NPI stages when Threshold NPI relaxation policy is applied

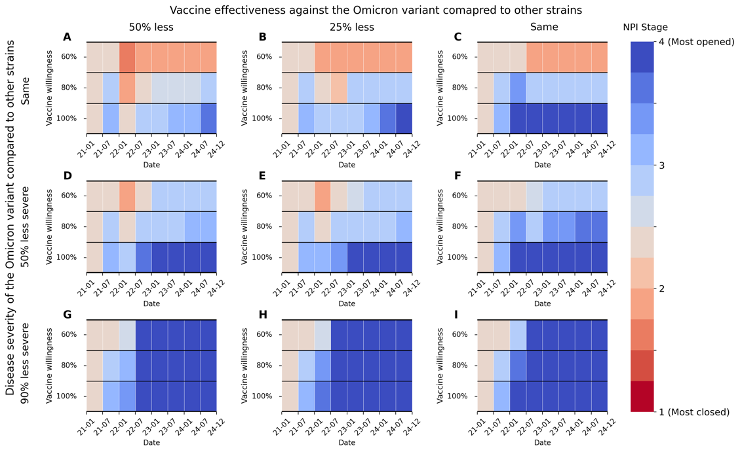

Figure 11 Changes in NPI stages (averaged by 6-month intervals) when Threshold NPI policy is applied with varying disease severity and vaccine effectiveness against the Omicron variant compared to other strains. Average immunity duration is 1 year and booster is scheduled for every 6 months.

#### 6.3. Detailed Results

Table 6 Detailed results when average immunity duration is 1 year and booster is scheduled for every 6 months. Scenarios that meet the objective on reducing annual death rate from COVID-19 at or below the level of influenza and pneumonia (12.6) are colored red. Scenarios that fully open the society from January 1, 2022 are filled yellow.

| Disease severity of the Omicron variant compared to other strains | Vaccine effectiveness against the Omicron variant compared to other strains | NPI policy | Vaccine willingness | Deaths during 2021-2024 | Annual Death Rate per 100 K during 2023-2024 |
| --- | --- | --- | --- | --- | --- |
| Same | 50% less effective | 1 | 60 | 10,183 | 118.8 |
|  |  |  | 80 | 6,561 | 71.7 |
|  |  |  | 100 | 3,906 | 18.5 |
|  |  | 2 | 60 | 7,004 | 96.2 |
|  |  |  | 80 | 4,460 | 40.3 |
|  |  |  | 100 | 2,567 | 5.8 |
|  |  | Threshold | 60 | 4,017 | 52.4 |
|  |  |  | 80 | 3,173 | 38.9 |
|  |  |  | 100 | 2,612 | 30.9 |
|  | 25% less effective | 1 | 60 | 9,493 | 121.1 |
|  |  |  | 80 | 5,517 | 68.3 |
|  |  |  | 100 | 2,390 | 10.0 |
|  |  | 2 | 60 | 6,325 | 95.1 |
|  |  |  | 80 | 3,525 | 38.6 |
|  |  |  | 100 | 1,449 | 4.0 |
|  |  | Threshold | 60 | 3,485 | 50.3 |
|  |  |  | 80 | 2,602 | 37.8 |
|  |  |  | 100 | 1,567 | 22.2 |
|  | Same | 1 | 60 | 7,943 | 120.1 |
|  |  |  | 80 | 3,427 | 52.6 |
|  |  |  | 100 | 210 | 1.7 |
|  |  | 2 | 60 | 4,809 | 80.2 |
|  |  |  | 80 | 1,788 | 28.3 |
|  |  |  | 100 | 9 | 0.0 |
|  |  | Threshold | 60 | 2,899 | 44.2 |
|  |  |  | 80 | 1,722 | 25.6 |
|  |  |  | 100 | 154 | 0.4 |
| 50% less severe | 50% less effective | 1 | 60 | 6,106 | 64.2 |
|  |  |  | 80 | 3,746 | 36.2 |
|  |  |  | 100 | 2,182 | 8.1 |
|  |  | 2 | 60 | 4,118 | 51.9 |
|  |  |  | 80 | 2,544 | 20.4 |
|  |  |  | 100 | 1,465 | 2.3 |
|  |  | Threshold | 60 | 3,223 | 39.5 |
|  |  |  | 80 | 2,834 | 37.2 |
|  |  |  | 100 | 1,889 | 18.9 |
|  | 25% less effective | 1 | 60 | 5,742 | 65.8 |
|  |  |  | 80 | 3,213 | 36.2 |
|  |  |  | 100 | 1,387 | 4.0 |
|  |  | 2 | 60 | 3,769 | 51.7 |
|  |  |  | 80 | 2,024 | 19.7 |
|  |  |  | 100 | 826 | 0.6 |
|  |  | Threshold | 60 | 2,886 | 38.8 |
|  |  |  | 80 | 2,378 | 33.7 |
|  |  |  | 100 | 1,192 | 14.5 |
|  | Same | 1 | 60 | 4,916 | 65.6 |
|  |  |  | 80 | 2,108 | 30.1 |
|  |  |  | 100 | 75 | 0.0 |
|  |  | 2 | 60 | 2,986 | 45.5 |
|  |  |  | 80 | 985 | 13.5 |
|  |  |  | 100 | 9 | 0.0 |
|  |  | Threshold | 60 | 2,590 | 37.2 |
|  |  |  | 80 | 1,483 | 19.5 |
|  |  |  | 100 | 141 | 0.0 |
| 90% less severe | 50% less effective | 1 | 60 | 2,587 | 18.3 |
|  |  |  | 80 | 1,346 | 8.1 |
|  |  |  | 100 | 741 | 0.2 |
|  |  | 2 | 60 | 1,534 | 13.1 |
|  |  |  | 80 | 858 | 4.8 |
|  |  |  | 100 | 519 | 0.0 |
|  |  | Threshold | 60 | 1,982 | 20.8 |
|  |  |  | 80 | 1,268 | 10.8 |
|  |  |  | 100 | 529 | 0.0 |
|  | 25% less effective | 1 | 60 | 2,454 | 18.5 |
|  |  |  | 80 | 1,176 | 8.3 |
|  |  |  | 100 | 502 | 0.0 |
|  |  | 2 | 60 | 1,435 | 13.1 |
|  |  |  | 80 | 704 | 5.0 |
|  |  |  | 100 | 309 | 0.0 |
|  |  | Threshold | 60 | 1,946 | 21.2 |
|  |  |  | 80 | 1,154 | 9.8 |
|  |  |  | 100 | 429 | 0.0 |
|  | Same | 1 | 60 | 2,218 | 19.1 |
|  |  |  | 80 | 745 | 5.4 |
|  |  |  | 100 | 69 | 0.0 |
|  |  | 2 | 60 | 1,200 | 11.8 |
|  |  |  | 80 | 296 | 2.3 |
|  |  |  | 100 | 9 | 0.0 |
|  |  | Threshold | 60 | 1,790 | 19.7 |
|  |  |  | 80 | 790 | 5.0 |
|  |  |  | 100 | 140 | 0.0 |

**References**

1.	Centers for Disease Control and Prevention. COVID-19 Pandemic Planning Scenarios – September 10, 2020. <https://www.cdc.gov/coronavirus/2019-ncov/hcp/planning-scenarios-archive/planning-ccenarios-2021-03-19.pdf>

2.	Campbell F, Archer B, Laurenson-Schafer H, et al. Increased transmissibility and global spread of SARS-CoV-2 variants of concern as at June 2021. *Eurosurveillance*. 2021;26(24):2100509.

3.	Centers for Disease Control and Prevention. COVID-19: When to Quarantine. 2021. [Online; accessed 18-July-2021]. <https://www.cdc.gov/coronavirus/2019-ncov/if-you-are-sick/quarantine.html>

4.	Institute for Health Metrics and Evaluation. COVID-19 model update: Omicron and waning immunity. 2021. [Online; Accessed 10-February-2022]. <https://www.healthdata.org/special-analysis/omicron-and-waning-immunity>

5.	Lauer SA, Grantz KH, Bi Q, et al. The incubation period of coronavirus disease 2019 (COVID-19) from publicly reported confirmed cases: estimation and application. *Annals of internal medicine*. 2020;172(9):577-582.

6.	Verity R, Okell LC, Dorigatti I, et al. Estimates of the severity of coronavirus disease 2019: a model-based analysis. *The Lancet Infectious Diseases*. 2020;doi:10.1016/S1473-3099(20)30243-7

7.	He X, Lau EHY, Wu P, et al. Temporal dynamics in viral shedding and transmissibility of COVID-19. *Nature medicine*. 2020;26(5):672-675.

8.	Young BE, Ong SWX, Kalimuddin S, et al. Epidemiologic features and clinical course of patients infected with SARS-CoV-2 in Singapore. *Jama*. 2020;323(15):1488-1494.

9.	Anderson RM, Heesterbeek H, Klinkenberg D, Hollingsworth TD. How will country-based mitigation measures influence the course of the COVID-19 epidemic? *The Lancet*. Mar 21 2020;395(10228):931-934. doi:10.1016/S0140-6736(20)30567-5

10.	Davies NG, Klepac P, Liu Y, et al. Age-dependent effects in the transmission and control of COVID-19 epidemics. *Nat Med*. Aug 2020;26(8):1205-1211. doi:10.1038/s41591-020-0962-9

11.	Centers for Disease Control and Prevention. COVID-19 Pandemic Planning Scenarios – May 20, 2020. <https://www.cdc.gov/coronavirus/2019-ncov/hcp/planning-scenarios-archive/planning-scenarios-2020-05-20.pdf>

12.	Dan JM, Mateus J, Kato Y, et al. Immunological memory to SARS-CoV-2 assessed for up to 8 months after infection. *Science*. Feb 5 2021;371(6529)doi:10.1126/science.abf4063

13.	Clark A, Jit M, Warren-Gash C, et al. How many are at increased risk of severe COVID-19 disease? Rapid global, regional and national estimates for 2020. *MedRxiv*. 2020;

14.	Makary M. Risk factors for COVID-19 mortality among privately insured patients. FAIR Health White Paper2020. [Online; Accessed 12-July-2021]. <https://s3.amazonaws.com/media2.fairhealth.org/whitepaper/asset/Risk%20Factors%20for%20COVID-19%20Mortality%20among%20Privately%20Insured%20Patients%20-%20A%20Claims%20Data%20Analysis%20-%20A%20FAIR%20Health%20White%20Paper.pdf>

15.	Murphy SL, Xu J, Kochanek KD, Arias E, Tejada-Vera B. Deaths: Final data for 2018. 2021. [Online; Accessed 12-July-2021]. <https://www.cdc.gov/nchs/data/nvsr/nvsr69/nvsr69-13-508.pdf>

16.	Holshue ML, DeBolt C, Lindquist S, et al. First case of 2019 novel coronavirus in the United States. *New England Journal of Medicine*. 2020;

17.	Port of Region A. Confirmed COVID-19 Positive Tests at SEA.

18.	Region A, Department of Health. COVID-19 Variants. 2021. [Online; Accessed 5-September-2021].

19.	Region A, Department of Health. Healthy Region A: Roadmap to Recovery Report. 2021. [Online; Accessed 18-July-2021].

20.	Eikenberry SE, Mancuso M, Iboi E, et al. To mask or not to mask: Modeling the potential for face mask use by the general public to curtail the COVID-19 pandemic. *Infectious Disease Modelling*. 2020;

21.	Region A- Department of Health. Case investigation and contact tracing dashboard.

22.	Kucharski AJ, Klepac P, Conlan A, et al. Effectiveness of isolation, testing, contact tracing and physical distancing on reducing transmission of SARS-CoV-2 in different settings. *medRxiv*. 2020;

23.	Lee S, Zabinsky ZB, Wasserheit JN, Kofsky SM, Liu S. COVID-19 Pandemic Response Simulation in a Large City: Impact of Nonpharmaceutical Interventions on Reopening Society. *Med Decis Making*. May 2021;41(4):419-429. doi:10.1177/0272989X211003081

24.	Region A, Department of Health. Interim COVID-19 Vaccination Plan-October 2020, version 1. 2020. [Online; Accessed June-1-2021].

25.	Region A, Department of Health. Region A’s COVID-19 Vaccine Phases. 2021. [Online; Accessed 1–June-2021].

26.	Food Drug Administration. Vaccines and related biological products advisory committee meeting december 10, 2020: FDA briefing document, Pfizer-BioNTech COVID-19 vaccine. 2020. [Online; Accessed 19-July-2021]. <https://www.fda.gov/media/144245/download>

27.	Thompson MG, Burgess JL, Naleway AL, et al. Interim estimates of vaccine effectiveness of BNT162b2 and mRNA-1273 COVID-19 vaccines in preventing SARS-CoV-2 infection among health care personnel, first responders, and other essential and frontline workers—eight US locations, December 2020–March 2021. *Morbidity and Mortality Weekly Report*. 2021;70(13):495.

28.	Business wire. Pfizer and BioNTech Confirm High Efficacy and No Serious Safety Concerns Through Up to Six Months Following Second Dose in Updated Topline Analysis of Landmark COVID-19 Vaccine Study. 2021. [Online; Accessed 31-May-2021]. <https://www.businesswire.com/news/home/20210401005365/en/>

29.	Region A - Department of Health. Summary of COVID vaccination among Region A residents.

30.	Region A, Department of Health. Everyone 12 and older now eligible for Pfizer-BioNTech COVID-19 vaccine. 2021.

31.	Stankiewicz K. Pfizer director Dr. Scott Gottlieb says Covid vaccine for kids 5 to 11 could come by winter. CNBC2021. [Online; Accessed 5-September-2021]. <https://www.cnbc.com/2021/08/30/pfizer-director-dr-scott-gottlieb-on-covid-vaccine-for-kids-5-to-11.html>

32.	Roy J. When will kids under 5 be able to get vaccinated for COVID-19? *Los Angeles Times*. December 17, 2021. <https://www.latimes.com/science/story/2021-12-17/whats-the-timeline-for-kids-under-5-to-get-a-covid-vaccine>

33.	IHME COVID-19 forecasting team. Modeling COVID-19 scenarios for the United States. *Nature medicine*. 2020;

34.	Region A Office of Superintendent of Public Instruction. School Facility ReOpening Survey.
